# Introducing a framework for within-host dynamics and mutations modelling of H5N1 influenza infection in humans

**DOI:** 10.1101/2024.09.01.24312235

**Authors:** Daniel Higgins, Joshua Looker, Robert Sunnucks, Jonathan Carruthers, Thomas Finnie, Matt J. Keeling, Edward M. Hill

## Abstract

Avian influenza A(H5N1) poses a public health risk due to its pandemic potential should the virus mutate to become human-to-human transmissible. To date, reported influenza A(H5N1) human cases have typically occurred in the lower respiratory tract with a high case fatality rate. There is prior evidence of some influenza A(H5N1) strains being a small number of amino acid mutations away from achieving droplet transmissibility, possibly allowing them to be spread between humans. We present a mechanistic within-host influenza A(H5N1) infection model, novel for its explicit consideration of the biological differences between the upper and lower respiratory tracts. We then estimate a distribution of viral lifespans and effective replication rates in human H5N1 influenza cases. By combining our within-host model with a viral mutation model, we determine the probability of an infected individual generating a droplet transmissible strain of influenza A(H5N1) through mutation. For three mutations, we found a peak probability of approximately 10^−3^ that a human case of H5N1 influenza produces at least one virion during the infectious period. Our findings provide insights into the risk of differing infectious pathways of influenza A(H5N1) (namely avian-human vs avian-mammal-human routes), demonstrating the three-mutation pathway being a cause of concern in human cases.

## 1 Introduction

The influenza virus family is responsible for influenza infections (colloquially referred to as the ‘flu’) in a variety of animals including humans, other mammals and birds. There are four main influenza types (A-D); within type A influenza there is substantial public health concern around the avian influenza A(H5N1) subtype, commonly known as bird flu. Influenza A(H5N1), which we will refer to as H5N1 influenza, is highly pathogenic in avian species and considered panzootic, being widely distributed in wild and domesticated birds [1]. As of 20 January 2025, worldwide there have been 964 reported cases of human H5N1 influenza and 466 deaths; a case fatality rate of approximately 50% reflects these reported cases having generally been severe [2].

At the time of writing, there is little evidence for human-to-human transmission of H5N1 [3]. Nonetheless, the high prevalence of the infection in the avian population is causing mounting concerns that under the right circumstances, an H5N1 strain could mutate to allow human-to-human transmission. If this were to occur, transmission between humans is likely to allow increased spread of the virus (at similar levels to the seasonal flu) with a resultant pandemic amongst humans.

Previous flu pandemics, and seasonal flu outbreaks, are primarily infections of the upper respiratory tract (URT) [4] due to the presence of *α*2,6-linked sialic acid receptors that these strains preferentially bind to for cell entry. H5N1 influenza, however, preferentially binds to *α*2,3-linked sialic acid receptors present in the avian respiratory and intestinal tracts [5–9], and these receptors are primarily found in the lower respiratory tract (LRT) in humans. This not only makes it much more difficult for initial human infection to occur, but also means that droplet transmission (the main source of seasonal flu transmission) is not viable, hence the current lack of human-to-human transmission of H5N1 influenza. However, with suitable mutations within humans, H5N1 influenza could evolve the ability to infect the URT as well as the LRT. This is cause for concern for two reasons. Firstly, infections in the LRT may lead to greater mortality due to increased risk of pneumonia and other related fatality risks [10]. Secondly, with the ability to infect the URT, human-to-human transmission becomes more likely, increasing the pandemic potential of H5N1 influenza [5, 7, 8].

Prior studies found that five amino acid substitutions in H5N1 influenza were required for human-to-human transmission to be possible at the time those experiments were conducted, with two of these mutations having already been seen in viruses sampled from the avian population [5, 7, 8]. It is believed that the other three mutations are unlikely to evolve in avian species as they are deleterious to the virus in birds [5]. Consequently, between three and five mutations are likely required to evolve within humans for an increased chance of droplet transmission.

For pandemic preparedness, it is crucial that we have suitable tools available to quantify the chance of an infected individual generating a droplet transmissible strain of H5N1 influenza through mutation. However, the probability of such mutations in H5N1 influenza occurring within a human host is presently unknown. To enable modelling analysis of this problem, there are two key limitations in the existing modelling literature. The first is that previous models of H5N1 influenza within-host infection dynamics in humans do not take into account the differences between the two tracts (URT and LRT). Although there have been modelling efforts to account for the binding specificities of H5N1 influenza in different areas of the respiratory tract [6], and it is understood that fluid dynamic effects/having multiple patches impact contagion dynamics [11–13], to our knowledge no current research explicitly models H5N1 influenza infection dynamics in the LRT and URT. The second is that although potential mathematical frameworks for the modelling of advantageous mutations (such as those required for droplet transmissibility) do exist in the literature, these have usually only explored the implications of the frameworks as opposed to explicitly finding the mutation probabilities [5, 14, 15]. For example, a study by Sigal *et al.* [16], to predict the fate of mutations that may arise *de novo* in the viral population, has presented a deterministic model of the within-host dynamics of early infection by a viral pathogen, coupled to a detailed life-history model using a branching process approach. That study, however, had a focus on seasonal influenza A viruses and was limited to exploring mutations affecting a single trait in isolation (i.e. it was assumed that it was not possible for multiple beneficial mutations to emerge and be transmitted to a new host).

In this paper, we present a combined modelling framework to address these two notable methodological gaps. The first modelling component is a novel within-host two-patch (both upper and lower respiratory tract), ODE infection model for H5N1 influenza. By inferring patch-dependent disease parameters, we seek to capture the biological differences in spreading capability of H5N1 influenza in the two parts of the respiratory tract. The second modelling component is an enhanced branching process model (BPM) for H5N1 influenza virus mutation, building on the work of Russell *et al.* [5]. Informed by the within-host model outputs, and including the distribution of infection lifespans and real-time replication number estimates, we use the BPM to provide a more realistic estimate on the evolutionary dynamics of a human H5N1 influenza infection. Combined, our modelling framework is generalisable to other respiratory pathogens, allowing researchers to estimate the mutation chances for a pathogen mutating specific traits.

## 2 Methods

Herein we summarise the three main methodological components of our study. We begin with a description of the novel within-host deterministic infection model and its calibration to both the canonical H5N1 influenza dataset and case fatality rate (Section 2.1). This is followed by the introduction of the branching process model for viral mutation and how it incorporated the within-host model results (Section 2.2). Finally, we list the methods and model realisations used to calculate both the time dependent proportion of mutant virions in a host and the probability that a human-to-human transmissible strain could arise from an infection (Section 2.3).

To simulate the within-host infection model (and its proxies) and the branching process model we used Python 3.11 with packages: Numpy (version 1.26.4), Matplotlib (version 3.84), Scipy (version 1.13.0) and Pickle (version 4.0). We conducted the Approximate Bayesian Computation scheme for fitting the within-host model in R 4.3 using the packages: tvmtnorm (version 1.6), KScorrect (version 1.4.0) and deSolve (version 1.40). A repository containing the data and code used to conduct this study can be found at https://github.com/joshlooks/avianflu.

### 2.1 Within-host infection model explanation and fitting

Our within-host model for H5N1 influenza infection introduced key biological processes not present in other models in the literature. This model development subsequently forms the basis of the remaining results presented in this paper. Here we outline the canonical dataset used for fitting the intra-host model (Section 2.1.1), provide the biological description of the infection model (Section 2.1.2) and state the corresponding ODE system (Section 2.1.3). We then explain how the model parameters were calibrated using literature (Section 2.1.4) and an Approximate Bayesian Computation scheme (Section 2.1.5). Lastly, introducing mortality into our two-patch model was of utmost importance to inform how likely human-to-human transmission may be. The relatively high case fatality rate of H5N1 influenza could hamper its ability to mutate in the body since those infected may be likely to die before the virus has a chance to mutate to become human-to-human transmissible. We thus conclude this section by outlining how we fit the model outputs to mortality data (Section 2.1.6). We also acknowledge that case fatality rate estimates for H5N1 influenza in humans are likely to be an overestimate of the true infection fatality rate because recorded cases have primarily been cases with severe symptoms requiring healthcare; we expand on the implications of this data caveat in the discussion.

#### 2.1.1 Data

We made use of the ubiquitous dataset in the literature corresponding to the viral titres of 18 hospitalised H5N1 influenza patients in Vietnam in 2004 and 2005 [17]. Of these 18 hospitalised patients, 13 died as a result of the H5N1 influenza infection episode. As the raw data were not publicly available, we estimated the viral titre values from Supplementary Figure 1 in de Jong *et al.* [17]; given the large variation in the data point values we expect our estimation procedure to have little effect on the takeaways presented. The titrations were formed from pharyngeal swabs taken after presentation at the hospital. These measurements corresponded to the viral load in the URT only, with viral loads varying in many orders of magnitude between patients on the same estimated day since onset of illness (Fig. 1).

**Fig. 1.**
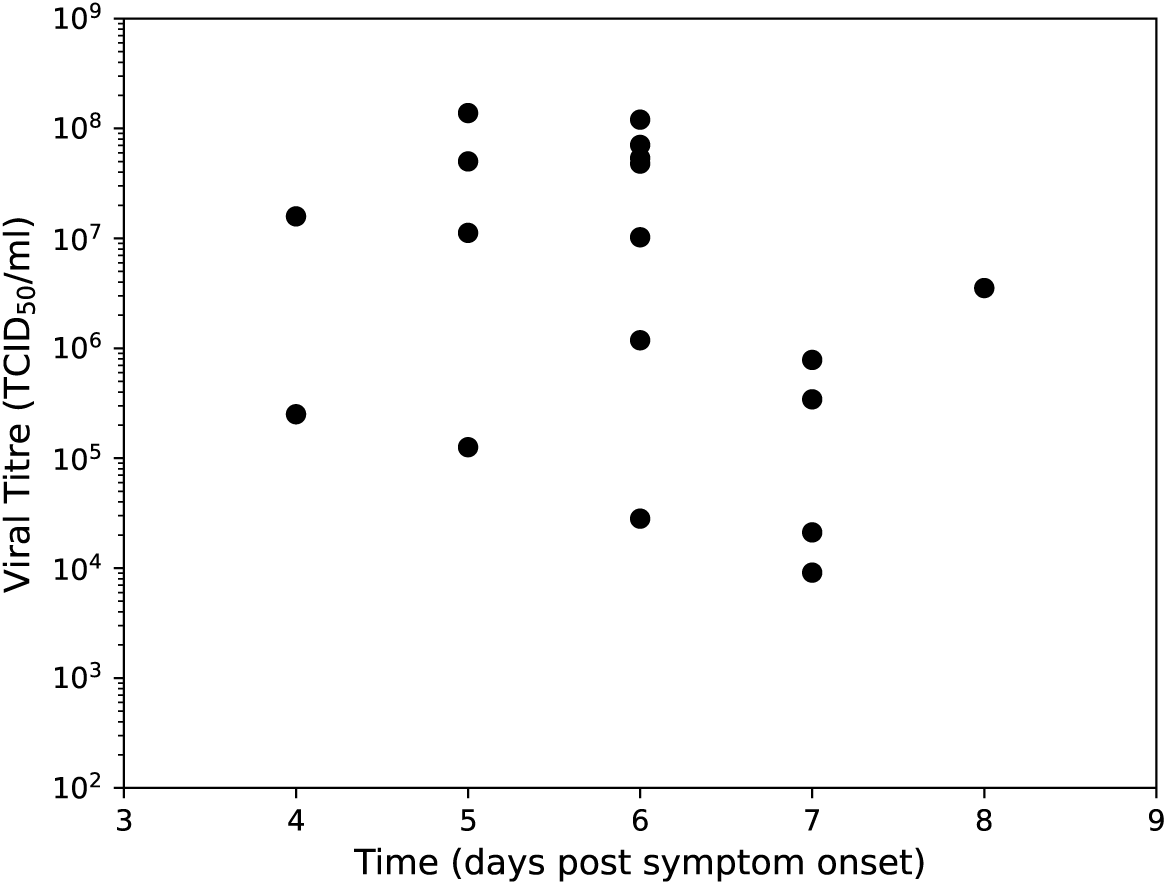
Viral titres from pharyngeal swabs of hospitalised H5N1 influenza patients in Vietnam. Data from de Jong *et al.* [17], where viral RNA loads were measured in throat swabs obtained at admission from 18 H5N1 patients. We used this dataset to calibrate all models included in this paper.

To calibrate our model output of days post-infection to the viral load data that was dependent on ‘days since onset of illness’, for our main analysis we implemented an incubation period of 0 days. This implementation matched an assumption applied previous modelling work by Dobrovolny *et al.* [6, 18] wherein they fit to the de Jong *et al.* [17] data models exploring the impacts of cell tropism and neuraminidase inhibitors on H5N1 infection. To assess the robustness of our findings to this implementation, we also performed our analysis assuming an incubation period of five days (i.e. model outputs on day 6 post-infection would correspond to day 1 since onset of illness); that estimate was informed by the median incubation period from a retrospective cohort study of 18 influenza virus (H5N1) cases in China [19]. We found similar qualitative findings when including the incubation period compared to when the incubation period was omitted (more details are given in Section S3).

We anticipated that this characteristic of the data, and the small, noisy sample, could pose issues around parameter identifiability and model generalisation to an ‘average infection’ during model fitting. Nonetheless, this dataset is the most recent and complete human infection data available for H5N1 influenza. Prior studies attempting to calibrate models to these data have gathered an understanding of related biological processes [6, 18]. It thereby provides an entry point for calibration of our proposed model and the exploration of its infection dynamics.

#### 2.1.2 Deterministic two-patch infection model description

We first built a deterministic two-patch ordinary differential equation infection model, with the URT and LRT each having their own internal processes. The URT and LRT then interact via the diffusion of the free virus between each patch and an advection term, describing the movement of free virus between patches via physical movement of fluid. The advection term can be considered the transfer of mucus (through coughing or mucociliary clearance by cilia) from the LRT to the URT. A graphical depiction of the above processes is shown in Fig. 2.

**Fig. 2.**
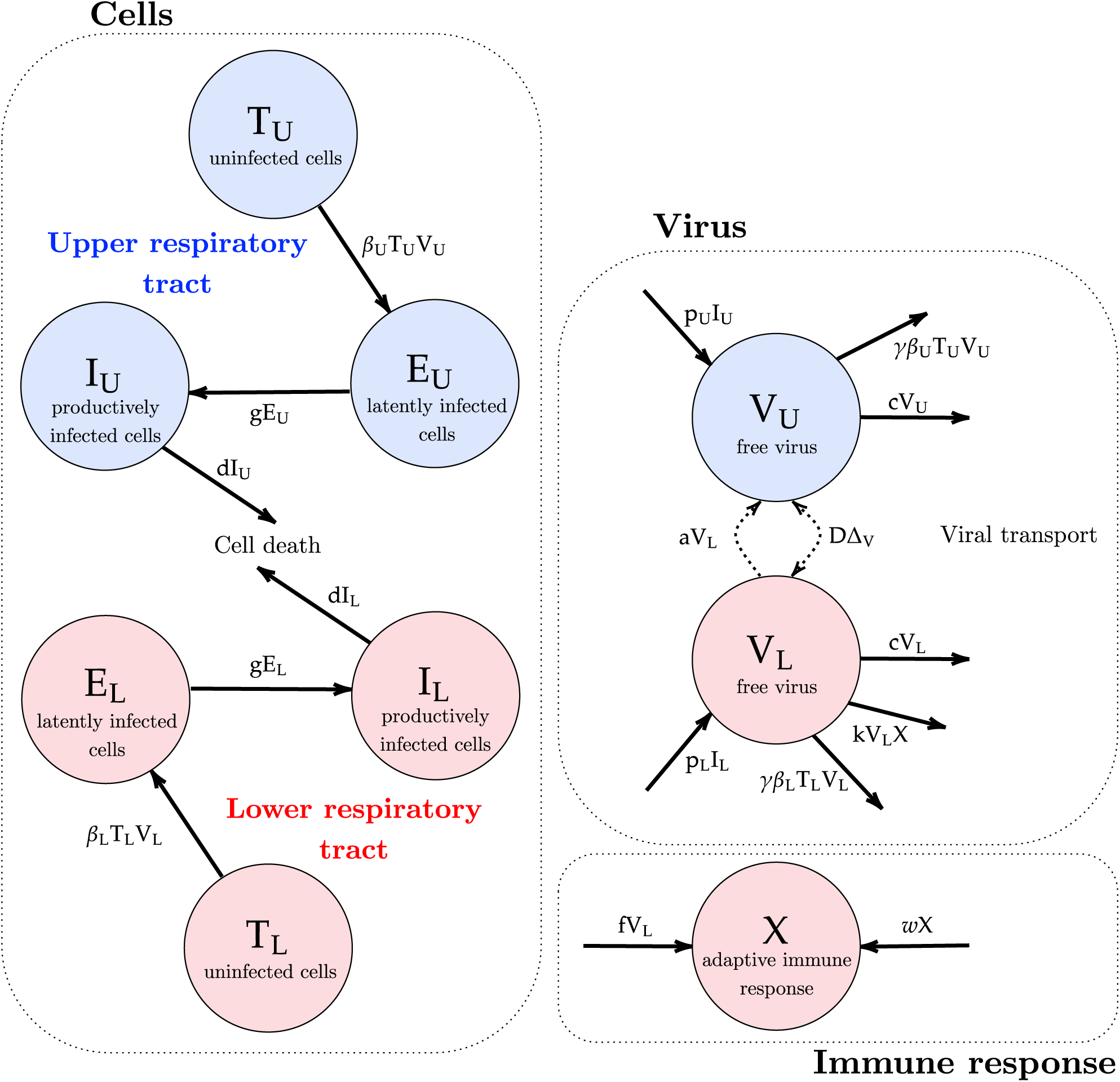
LRT and URT explicit within-host respiratory infection model schematic. Compartments listed are uninfected/target cells (*T*), free virions (*V*), eclipse/latent cells (*E*), infected/virion-producing cells (*I*) and the adaptive immune response (*X*). Note that the subscripts *U, L* represent the URT- and LRT-based compartments respectively. The different colours represent the processes in the URT (in blue) and in the LRT (red). Arrows show the spread of the contagion through the host. The dashed arrows in the virus compartment indicate the coupling of the two patches through advection and diffusion. Parameters descriptions are found in Table 1.

**Table 1.**
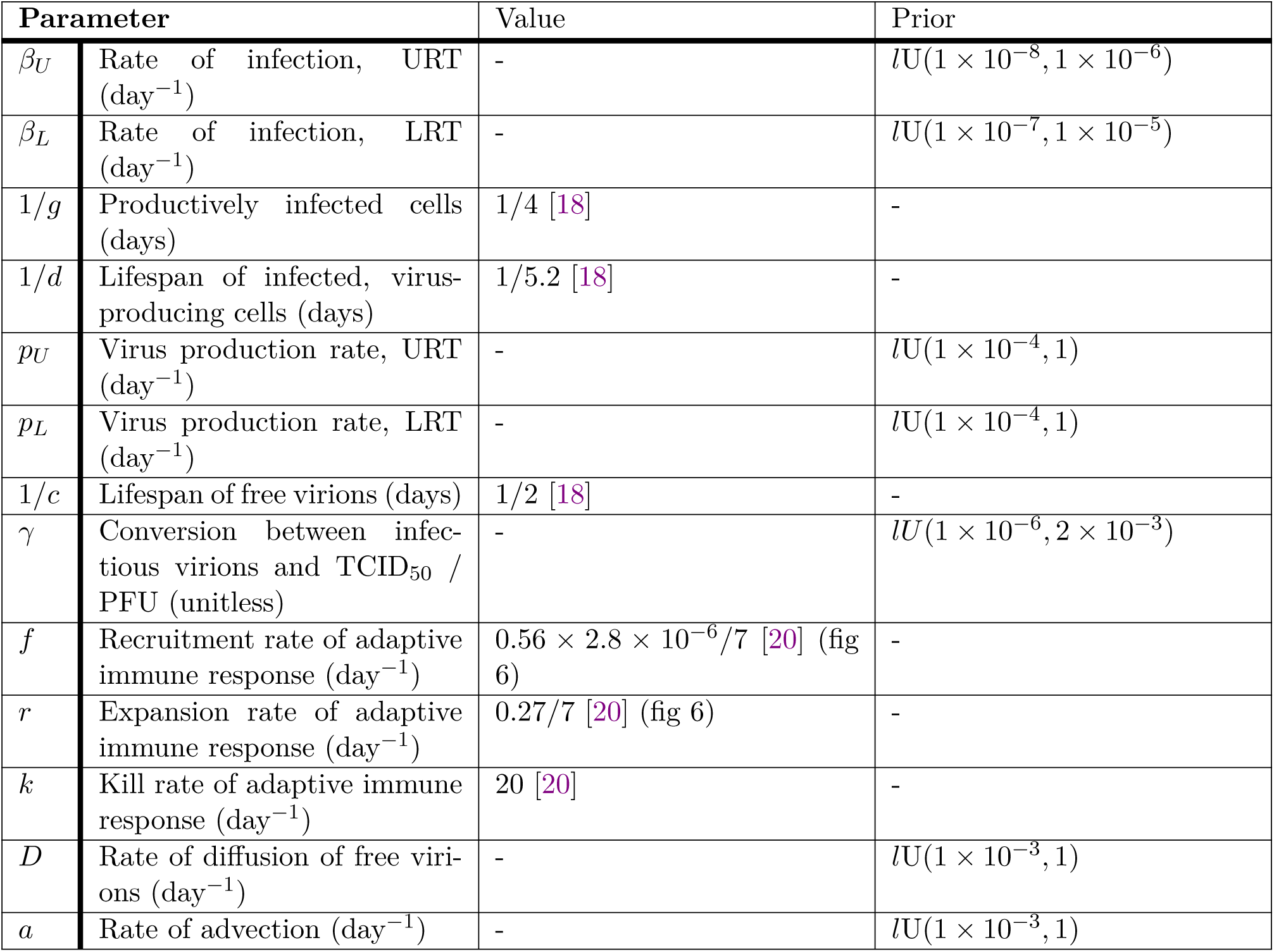
List of model parameters and their descriptions. For fixed parameters we state their value and associated references. For inferred parameters we list their prior distribution (we use *lU* as a notation for the log-uniform distribution). We provide unit information for each parameter in parenthesis after the parameter description.

For the within-patch processes (the cells subfigure in Fig. 2), we modelled each respiratory tract compartment as having a set of uninfected epithelial cells (or ‘target cells’, *T*) to which the H5N1 influenza virions (*V*) may bind. After infection by a virion, the cells move into an eclipse/latent phase (*E*) where they are infected by the virus but do not produce any additional virions. After an exponentially-distributed period of time, the cells leave the latent phase and enter the infected phase (*I*), producing free virions. LRT models for Influenza A have been studied previously; we based our more complex two-patch model on a model of the infection in the LRT by Handel *et al.* [20]. We note in particular that the key difference between the URT patch and the LRT patch is that it is generally considered that the URT can be modelled using a ‘target-cell limited’ approach. In other words, there is limited immune response in the URT and the dynamics of the virus are entirely governed by the number of uninfected cells alive. Thus, we only considered an immune response in the LRT patch. The adaptive immune response (*X*) has a humoral component comprised of B-cells and antibodies, as well as a cellular component, comprised of T-cells. The humoral component causes the IR to increase proportionally to the viral load in the LRT, and the clonal expansion of the T-cells causes the IR to grow exponentially, as in Handel *et al.* [20]. *X* can be considered to represent antibodies in the host.

#### 2.1.3 ODE system

The within-host dynamics of H5N1 infection obeyed the following system of ordinary differential equations. We note that a subscript *U* denotes that the compartment / parameter is for the URT, while a subscript *L* denotes that the compartment / parameter is for the LRT.

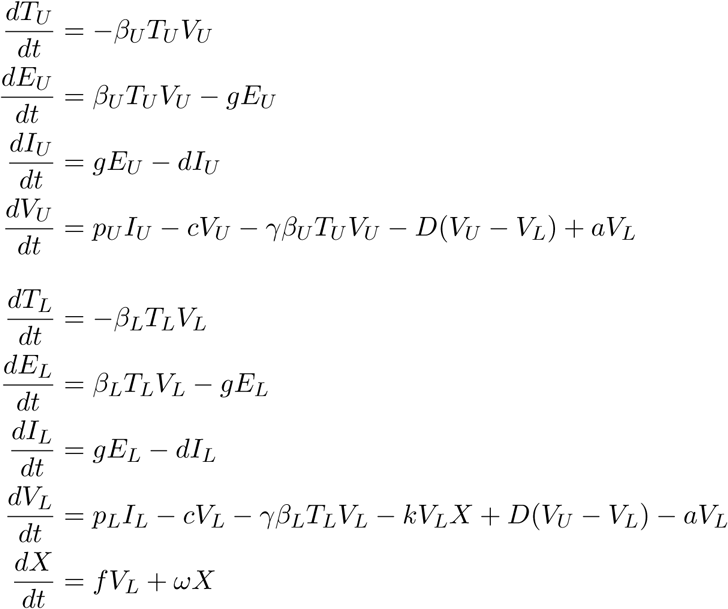

with *β_U_*and *β_L_* the rate of infection in the URT and LRT, *g* the latent transition rate of infected cells, *d* the mortality rate of infected virus producing cells , *p_U_*and *p_L_* the virus production rate in the URT and LRT, *c* the morality rate of free virions, *γ* the conversion rate between infection and viral titre (also referred to as the viral uptake rate), *f* the recruitment rate of adaptive immune response, *ω* the expansion rate of adaptive immune response and *k* the kill rate of adaptive immune response, *D* the rate of diffusion and *a* the rate of advection.

#### 2.1.4 Model parameterisation from the literature

We obtained values from the literature for a subset of parameters in our ODE model. Specifically, we used the parameter point estimates reported by Dobrovolny *et al.* [18] to set the latent state duration of infected cells (1*/g*) as 1*/*4 days, the lifespan of infected virus producing cells (1*/d*) to be 1/5.2 days and the lifespan of free virions (1*/c*) as 1/2 day. We also highlight that Dobrovolny *et al.* [18] noted that their values were consistent with other research in the area.

For the immune parameters, we took the approach found in Handel *et al.* [20]. Although this was fitted to mice data, studies have shown that the mice immune system is a suitable analogue for the immune system found in humans *in vivo* [21]. Further, parameters are likely transferable through the comparison of mice and human metabolic rates - mice have a metabolic rate seven times that of humans [22]. We converted from plaque-forming unit (pfu) into TCID_50_ (Tissue culture infectious dose 50%), with pfu being proportional to TCID50 by a factor of 0.56 [23].

It was also important to select an initial number of target cells and initial viral load. We took the estimated values of *T_U_* = 4 × 10^8^*, T_L_* = 6.25 × 10^9^ from Ciupe and Tuncer [12], which were calculated using the average surface area of an epithelial cell and of the human respiratory tract. We took the initial viral load (*V*_0_ = 1.3 × 10^3^ TCID_50_/ml) from the fitted values of the single-target-cell model in Dobrovolny *et al.* [6].

There was little information in the literature regarding rate of infection *β_U_* and *β_L_*, virus production rate *p_U_* and *p_L_*, conversion rate (rate of viral uptake) between infection and viral titre *γ*, rate of diffusion *D* or rate of advection *a*. These parameters of interest were also chosen as they have been found to have the biggest impact on the observed disease dynamics [6, 13, 18]. It is also worth noting that setting *γ* = 0 leads to similar results (and is normally ignored in human models [13, 18, 20]). This parameter represents the uptake rate between the viral titre (in TCID_50_) and the number of free virions used to infect a target-cell. Setting this parameter to zero indicates that there is no noticeable change in the viral titre due to the infection of target-cells. By re-introducing this parameter (allowing it to be non-zero), we gained an extra degree of freedom in the model that allowed for more biologically realistic parameter values and peak shapes to be observed during parameter fitting.

We state the default model parameters, for non-fitted parameters, in Table 1. Some of the selected parameter values are similar to literature values for models fitted to H1N1 infection data within humans [24–26]. However, previous studies on H5N1 influenza infection in humans found that these values gave good fits to the data, and that the other aforementioned parameters that we fitted for were the main contributors to viral dynamics [6, 18].

#### 2.1.5 Model calibration and parameter inference

To calibrate the model, we made use of the dataset outlined in Section 2.1.1. We note that these data correspond to the viral load in the URT only, meaning we could only fit the model dynamics based on the URT compartments. Parameter identifiability is a problem for most mathematical biology models, and this was especially true for our fitting process as we had less than 20 data points available, all of which corresponded to hospitalised individuals who died from the infection.

To fit the parameters we employed an Approximate Bayesian Computation Sequential Monte Carlo *M* Nearest Neighbours (ABC-SMC-MNN) method based on the pseudo-code found in Minter and Retkute [27], using methods originally developed by Filippi *et al.* [28] and Toni *et al.* [29]. Due to the lack of data, and its continuous nature, an exact likelihood function for data fitting is difficult to justify, thus we adopted an ABC inference scheme. With large order of magnitude differences across our data points, we chose the summary statistic (*c*) to be the model error on a log-scale, where *y* is the data, *N* is the number of data points and *x* is the model predictions:

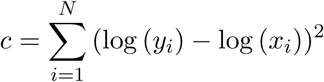

We chose the perturbation kernel to be a truncated-multivariate-normal distribution (truncated to take into account the prior). For the prior distributions, we assumed log-uniform prior distribution for all variables (see Table 1 for the prior distribution ranges). We selected log-uniform priors as it is an uninformative prior and because the parameters were likely to be skewed towards lower orders of magnitude (such that our prior belief was the parameters being uniform on a log-scale). We informed the parameter ranges of the priors by first taking a least-squares fit (to both the normal and log-scale data); we then took a wide range around those values to define the prior bounds. Furthermore, we also assumed the spreading rate in the LRT (*β_L_*) to be greater than that in the URT (*β_U_*). This is because H5N1 influenza preferentially binds to proteins more commonly found in the LRT as the type of receptor expression in the LRT is more similar to the avian respiratory tract [6, 30–32]. The chosen hyper-parameters for the algorithm were to run the method adaptively, with an error tolerance in the first generation of 180. The error tolerance for subsequent generations was then set at the 40^th^ percentile of the previous generations’ particles. We set the algorithm to terminate either after 10 generations, or when the error tolerance changed by less than 1% between subsequent generations. We ran the algorithm for the full ten generations, with a final (adaptive) error threshold of 141.58 (compared to a 142.1 tolerance in the previous generation). The error threshold in the final generation had similar error to the least-squares fit value of 120.9.

#### 2.1.6 Mortality

It is currently believed that a leading cause of death amongst H5N1 influenza patients is a phenomenon known as a ‘cytokine storm’ [33]. This occurs when the immune response to the virus is elevated to the point where the body overwhelms itself, causing massive inflammation and ultimately death [34].

Since a cytokine storm results from the immune system’s sustained response to viral load, for our two-patch model we took cumulative viral load as a proxy for mortality. In particular, we considered the integral of the logarithm of the viral load over time as our metric for mortality:

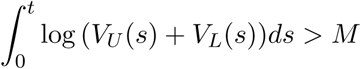

with *M* a constant. Although the immune response in the LRT may be more closely related to H5N1 fatalities (and so just using the viral load in the LRT may also be a reasonable proxy), viral load in the URT is still likely to affect mortality and so we assume that the cumulative viral load is a reasonable proxy for mortality. Further, we expect there to be more virions produced in the LRT (due to the larger number of cells) than the URT, meaning it should also contribute more to mortality. To determine the value of *M* , to represent a worst case scenario we took a case fatality rate that matched the mortality rate from the hospital patient cohort in the de Jong *et al.* [17] dataset (approximately 73%; 13 of the 18 patients). We then found the value of *M* that corresponded to said case fatality rate from the results of our stochastic simulations. In doing so, we set *M* = 154.760.

We also conducted a sensitivity analysis of our results to a lower case fatality rate of 20%. This value was taken from Dobrovolny *et al.* [18] for individuals treated with neuraminidase inhibitors and led to a higher *M* value of 177.328.

Introducing the proxy for mortality given above, we calculated the total length of infection for each of our infection simulations. We considered the infection to be finished when either a patient dies or their total viral load fell below *V_U_* + *V_L_ <* 10^4^ (a threshold of 10^4^ total virions was chosen to be two orders of magnitude less than our estimate of an infection inoculum size of 10^6^ virions – see Section 2.3 for our heuristic derivation of the infection inoculum size), i.e.

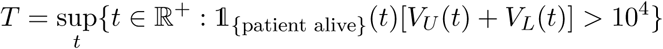

From this, we calculated an empirical distribution for *T* that we used to model viral mutations within humans (Fig. 3(a)). When taking a case fatality rate of approximately 73%, most of the empirical distribution for *T* occurred between eleven and a half to twelve and a half days post infection, having reasonable correspondence to previously recorded infections of (and modelling efforts for) H5N1 influenza infections lasting for around ten days [5, 6, 18]. When instead assuming a case fatality rate of 20%, most of the empirical distribution for *T* was between twelve and fifteen days (Fig. 3(b)).

**Fig. 3.**
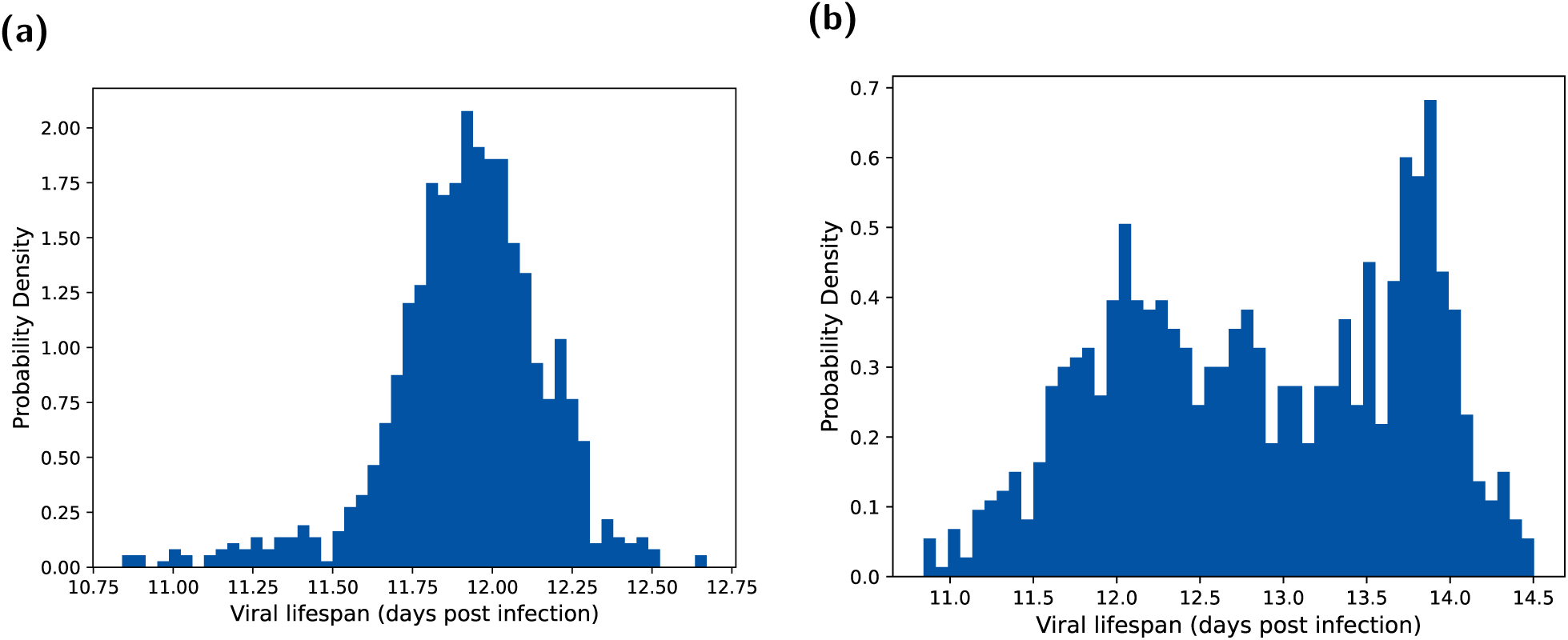
Viral lifespan (*T*) distributions under each case fatality rate assumption. The assumed case fatality rates were **(a)** 73% and **(b)** 20%, respectively. We obtained the viral lifespan distributions by determining when either the viral load dropped below 10^4^ or the integral under the log curve reached a value *M*. We performed the fitting method outlined in Section 2.1.6.

### 2.2 Mutation modelling and viral dynamics

From outputs that could be generated from our two-patch within-host model, we next needed an additional modelling component that would enable us to calculate the proportion of virions with zero, one, two, three, four and five mutations, and the probability that any given virion within the body had this number of mutations. In this section we outline our adapted stochastic branching process mutation model used for this purpose. This model contains biologically informed values for key model parameters, informed by the incorporation of results from the within-host infection model, thus providing a prominent modelling advance.

We adapted the stochastic branching process mutation model for viral mutation introduced in Russell *et al.* [5], in which viral replication occurs at fixed time intervals of length Δ with a mutation rate *µ* and replication rate *r*. The total number of viruses with *j* mutations at each time step *t_k_* = *k*Δ (with *k* ∈ N and *t_k_ < T*), 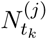, is then given as a Poisson random variable:

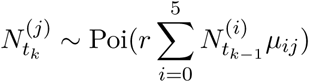

Where

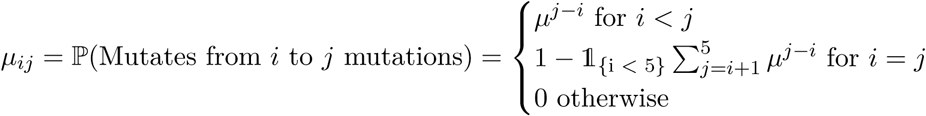

Note that the rate of 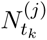 is a summation due to the additive property of the Poisson random variable. We then modified the above process to allow model parameters to be informed from the fitted within-host infection model. We allowed *r* to be a function of time, *r*(*t_k_*), rather than a fixed value. Our branching process was thereby defined by:

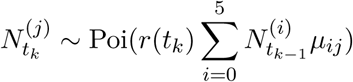

The function *r*(*t_k_*) represents the viral replication rate as derived from our two-patch within-host model. To calculate this, we need the growth rate of new virions and the death rate of existing virions.

We first express a partition *P* of [*t_k_, t_k_*_+1_] such that *t_k_* = *τ*_0_ *< τ*_1_ *<* · · · *< τ_m_* = *t_k_*_+1_ with *τ_i_*_+1_ − *τ_i_* = *δ* where *δ* is the rate at which the ODE system is updated when solved numerically.

Taking *S* ∈ {*U, L*}, to denote whether the value corresponds to the URT or LRT, and *N, V ^U^ , p, I, c, V ^L^, β, T, k, X* and *V* to be as described in Section 2.1.3, we now define the growth rate of new virions at time *τ_l_*:

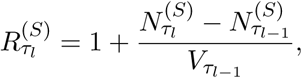

where *N* ^(*S*)^ solves the following differential equation for the rate of virion production:

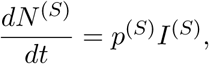

and we define the death rate of existing virions at time *τ_l_*:

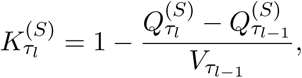

where *Q*^(*S*)^ solves the following differential equation for the rate of virion removal

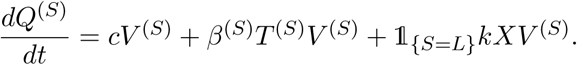

Our viral repilication rate *r*(*t_k_*) is then given by the product of the weighted sum of the number of virions created and destroyed at each time step *δ* in each tract:

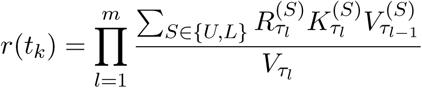

### 2.3 Mutant virion proportions and probabilities

Our final piece of analysis involved exploring the time dependent proportion of mutant virions in a host and the probability that a human-to-human transmissible strain could arise from an infection. Previous studies have found that up to two of the required five H5N1 influenza mutations can naturally occur in birds [5]. Depending on the number of mutations that have occurred prior to the human H5N1 influenza case, mutant virions then require either three out of three, four out of four, or five out of five of the required mutations for droplet transmission. Results from the branching process model allowed us to inform the probability that at any given time during the infection, the human host has at least one virion with the necessary number of mutations required for human-to-human transmission. Note that we term ”X out of X mutations” for instances where the required total five mutations to achieve droplet transmission could be obtained during the infection episode of the human case (i.e. acquiring three or more mutations during the human infection case episode).

We ran the branching process model for viral mutation over the 1000 posterior predictive trajectories acquired via the procedure outlined in Section 2.1.5. We initialised the starting viral load as 10^6^ virions in each realisation. Our reasoning for that choice is as follows. The initial viral count in our two-patch within-host model was 1.3 × 10^3^ TCID_50_/ml. For influenza A virions, the viral count per ml is around four orders of magnitude greater than the TCID_50_/ml value [35]. Using these two pieces of information, this gave us a viral density of approximately 10^7^ virions per ml. Then, taking an initial infected droplet of size 10^−1^ml, we arrive at an initial viral count of 10^6^ virions. In these simulations we also took Δ (the period between replications) to be six hours, noting that *δ* (the update rate of the two-patch within-host model solutions) is 0.001 days. This corresponds to the virions making two replication cycles (one from cRNA to vRNA and then back to cRNA) every 0.5 days, as in Russell *et al.* [5].

We ran two sets of simulations of the branching process model. The first was a set of one million BPM realisations (1000 copies of each of the 1000 sets of parameter samples in the posterior distribution), seeding the infection with an initial viral load of 10^6^ virions.

The second was a set of 1000 BPM realisations (one for each of the parameter sample sets in the parameter posterior distribution) with 10^6^ × 10^6^ initial virions (to simulate one billion people, but combining BPMs to save on computation time). This provided a higher precision in the calculation of mutation virus proportions.

We also calculated the probability that an individual had at least one virion exhibiting a specific number of mutations. This provided another indication of the likelihood of an infection mutating to allow for human-to-human droplet transmission. This probability calculation was, however, intractable for the BPM that simulated one billion people as it required being able to differentiate between individuals (not possible here as we combined BPMs as it is computationally expensive to run the number of individual realisations needed to achieve the required level of precision). We thus introduced an upper-bound estimate for this probability at time *t*. Using this approximation allowed for a probability approximation to be produced for a much higher number of BPM realisations.

The approximation was as follows. Let *V_t_* be the (mean) virion count for an individual at time *t*, and 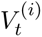 be the (mean) number of virions with *i* mutations for an individual at time *t*. Additionally, given an (average) infected individual, let *A* be the event that this individual has no virions with *i* mutations at time *t* and *B_k_*be the event that virion *k* in this individual does not have *i* mutations at time *t*, where *k* = 1, 2, …, *V_t_*. Then, Π(An individual at time *t* has at least one virion which has undergone *i* mutations)

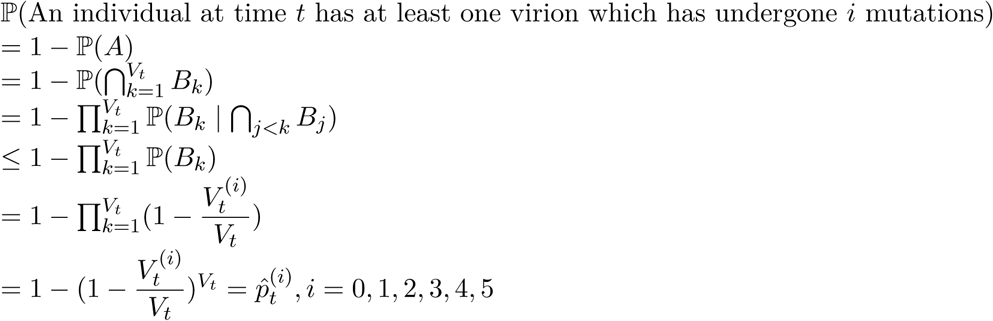

To justify the inequality, we first note that if an arbitrary virion at the current timestep has *i* mutations, the probability that any other virion has that number of mutations would increase. This is because there is a chance that virions with the same number of mutations could have the same parent. The joint probability events 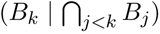 takes into account this positive correlation (but the event *B_k_* by itself does not), i.e. 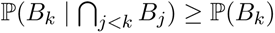

Additionally, since each virion is equally likely to mutate, we used the proportion of virions with *i* mutations to get that 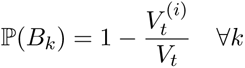.

## 3. Results

### 3.1 Fitting the two-tract within-host respiratory infection model to H5N1 influenza viral titre data

Having developed our within-host respiratory infection model, with infection dynamics in the LRT and URT modelled explicitly, it was important to ascertain whether it could reproduce the observed H5N1 influenza viral titres (Fig. 1) whilst maintaining biologically reasonable parameters. Resultant parameter posteriors would then be used as inputs to the branching process model.

We ran the ABC-SMC-MNN routine and obtained 1000 samples of the posterior distribution for seven fitted parameters: *β_U_* , *β_L_*, *p_U_* , *p_L_*, *γ*, *D* and *a* (Fig. 4). We note that even for the posteriors that had similarities to a log-uniform distribution (*β_L_, p_L_, γ, D, a*), the range and probability mass of these distributions shifted compared to the prior. This is reinforced by a least-squares fit producing a similar profile to the median of the posterior-predictive distribution (Fig. 5(a)). The least-squares fit parameters can be found in Section S1.

**Fig. 4.**
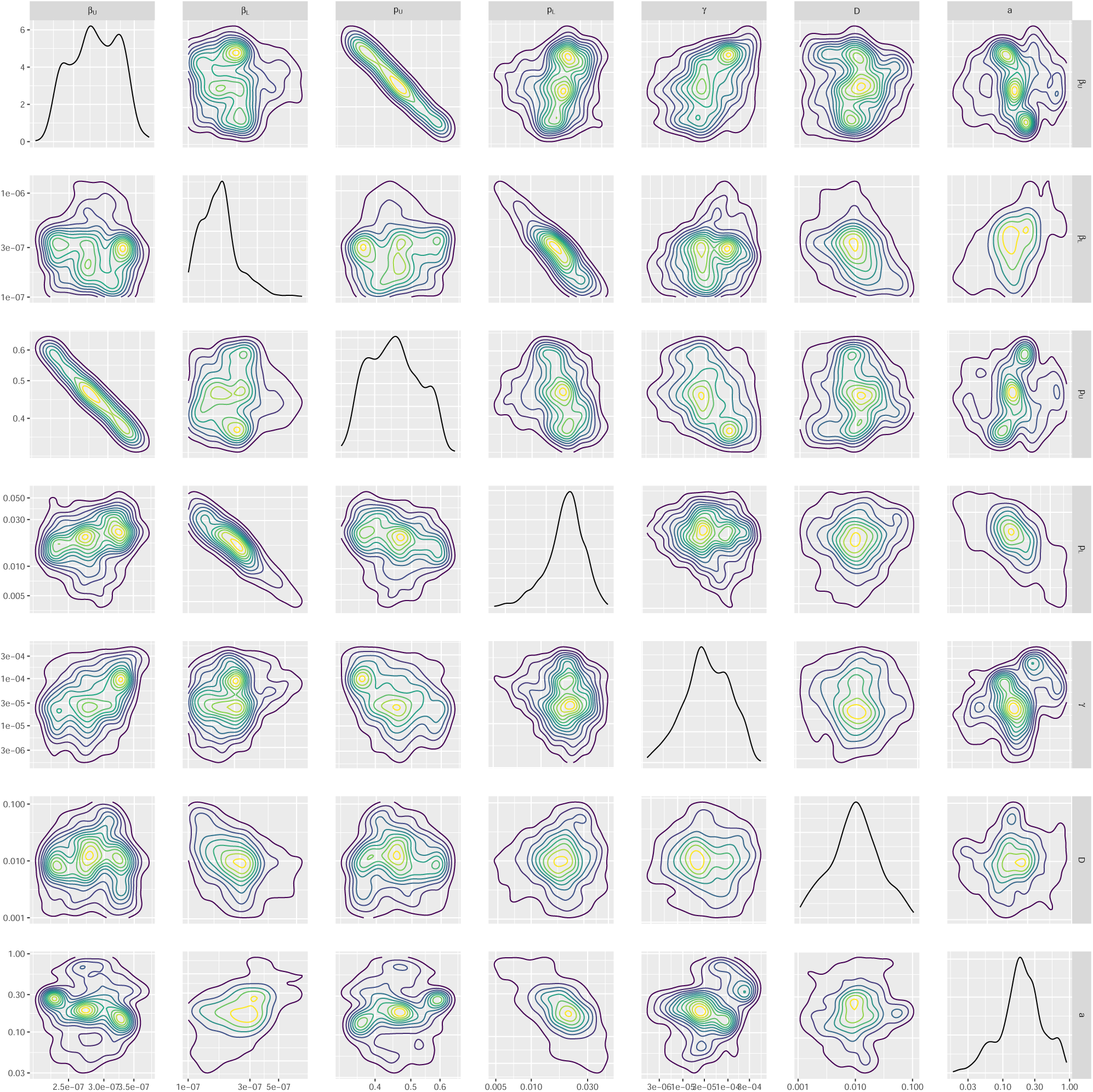
Parameter posterior distributions. We obtained 1000 samples of the target posterior distribution using the ABC-SMC-MNN method outlined in Section 2.1.5. Diagonal panels show the marginal distributions for: rate of infection in the URT (*β_U_*) and the LRT (*β_L_*), virus reproduction rate in the URT (*p_U_*) and the LRT (*p_L_*), viral uptake rate between infection and viral titre (*γ*), rate of diffusion of free virions (*D*) and the rate of advection (*a*), respectively. Off-diagonal panels show bi-parameter distributions, where the contour shading intensity corresponds to the probability density value (lighter for higher probability density). Parameters (*β_L_, p_L_*) in the LRT tended to be higher than the URT (*β_U_ , p_U_*), agreeing with the biological preference for H5N1 influenza to infect the LRT.

**Fig. 5.**
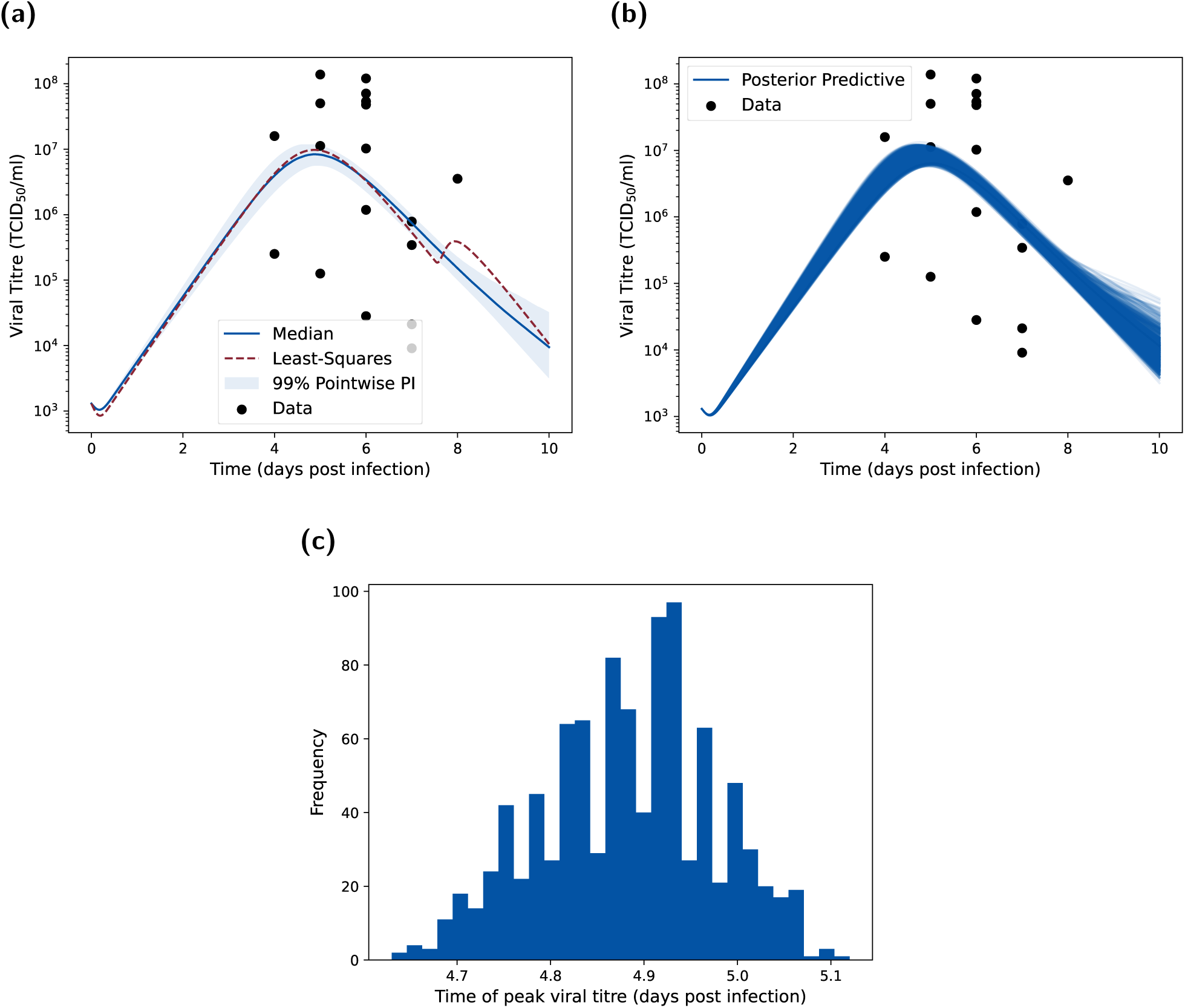
Posterior predictive distributions for URT viral titre metrics. **(a)** Posterior predictive distribution for *V_U_* compared to the empirical data. We produced the posterior predictive distribution using the 1000 parameter samples from our inferred parameter posterior distribution in Fig. 4. We display the median (blue solid line), 95% pointwise prediction interval (shaded region) and the least-squares fit (dotted red line). Both the optimisation fit and ABC posterior show reasonable concordance to the main data trends. **(b)** As for **(a)**, but showing all posterior predictive trajectories as opposed to the distribution summary. **(c)** Posterior viral titre peak-time distribution showing that the majority of infections peak just before day five. This is consistent with the data, which shows a peak around day five to day six (post infection).

Comparing the inferred posterior distributions for the URT and LRT spreading rate parameters, the 95% credible interval for the spreading rate in the URT (*β_U_* ∈ [2.24 × 10^−7^, 3.47 × 10^−7^]) was generally at a lower range than in the LRT (*β_L_*∈ [1.09×10^−7^, 1.18×10^−6^]). This difference possibly corresponds to the preferential binding of H5N1 influenza to the epithelial cells in the LRT than in the URT. The production rate in the URT (*p_U_* ∈ [0.348, 0.608]) was higher than in the LRT (*p_L_* ∈ [0.00488, 0.044]), likely due to the higher target-cell count (and thus maximum production rate) in the LRT. There was a clear negative correlation between *β_U_* and *p_U_* (relating to the previous discussion), which is to be expected as an increase in the spreading rate would lead to target-cells being infected sooner and hence a larger infection time available to produce virions (meaning that a lower *p_U_* is required) and vice-versa. The 95% credible interval for *γ* was at a low range of [2.72 × 10^−6^, 2.97 × 10^−4^], indicating that the parameter was needed to delay the peak time, but only at smaller values. The 95% credible intervals for the diffusion (*D* ∈ [0.00141, 0.0680]) and advection (*a* ∈ [0.0414, 0.731]) coefficients are quite wide, possibly indicating that the intra-patch processes contribute more than the inter-patch processes to the viral dynamics.

Through simulation of our model using the 1000 parameter sets representing samples from the target posterior distribution, we next checked the correspondence between the posterior predictive distribution for *V_U_* and the empirical viral titre data (Fig. 5(a)). As the data is wide ranging - the smallest viral titre measured had a value of around 10^4^*TCID*_50_*/ml* with the highest being at over 10^8^ - the likelihood surface has many steep local minima giving a tight posterior interval. However, it can also be seen that the least squares solution (Section S1) lies close to the median produced by the ABC method, giving confidence about the validity of the solution. The predictive interval lay within the middle range of the dataset. The qualitative shape (including peak height and time) of the median was very similar to other models [6, 13, 18, 36]. For the least-squares optimisation we chose five different starting parameter sets and selected the resultant local mode with the lower error. Although not guaranteed, we are confident that this is likely the global optima as multiple starting points converged to this value. This outcome supports the parameter posterior distributions acquired by the ABC approach successfully incorporating the posterior.

Lastly, inspection of the peak time distribution of viral titre realisations from the posterior predictive distribution showed almost all of the density of peak viral titre occurring between 4.7 and 5 days post infection (Fig. 5(c)). This observation provided further assurance in the concordance between the fitted model and the empirical data.

### 3.2 Viral dynamics and branching factor by survival status

Having acquired posterior predictive trajectories for the viral load, we fit the resultant values using the mortality proxy (Section 2.1.6). This process allowed for the separation of simulated stochastic viral dynamics into individual who cleared the infection and those who died (Fig. 6(a)).

**Fig. 6.**
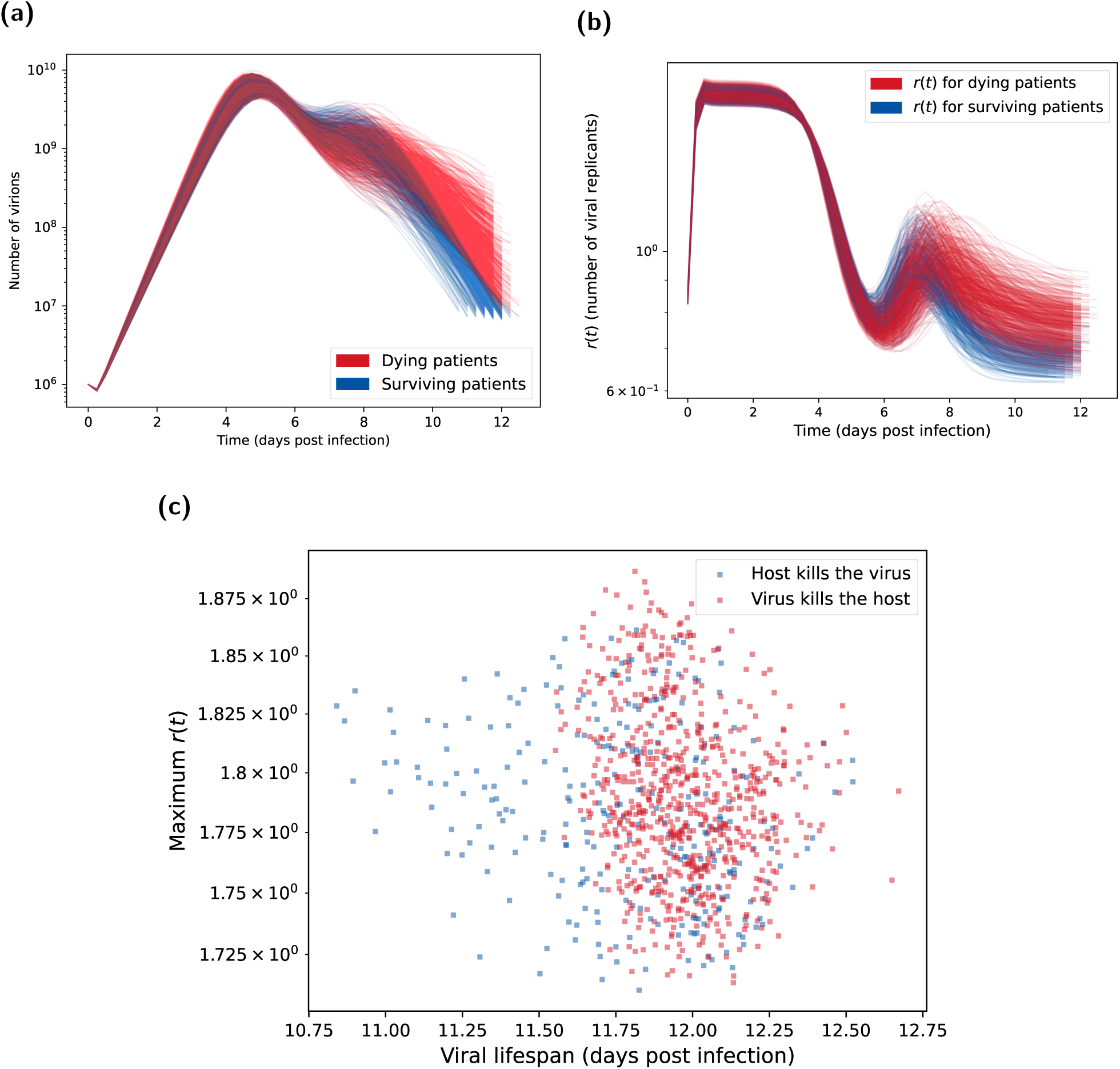
Posterior predictions for number of virions and number of viral replicants (*r*(*t*). Both plots show the 1000 posterior trajectories, with the blue lines representing H5N1 influenza patients who survive the infection (cleared the virus) and the red lines representing patients who died due to the infection (where the distinction is made using the method in Section 2.1.6). **(a)** Virion count distribution found using the parameter posterior in Fig. 4. After day 6, the viral count trajectories for deceased patients are higher and more sustained than surviving patients. These were calculated from the one million BPM realisations each with an initial viral load of 10^6^ and a mutation chance of 10^−5^ per replication. **(b)** Distribution of number of viral replicants (*r*(*t*)) calculated from the posterior predictive distribution shown in Fig. 5(a). Surviving patients tended to have higher *r*(*t*) during the second peak around day 7, which then declined below one (indicating a decreasing virion count) more rapidly than for dying patients. **(c)** Maximum ***r*** value vs viral lifespan. Maximum *r* values taken from Fig. 6(b) and corresponding viral lifespans are shown in Fig. 3. Blue circles represent H5N1 influenza patients who survive the infection (cleared the virus). Red circles represent patients who died due to the infection (where the distinction is made using the method in Section 2.1.6). Surviving individuals tended to have shorter viral lifespans. Otherwise, there was no evident dependencies between maximum *r* value and the viral lifespan.

Noting that the virion count is proportional to the viral titre (and so should follow the same dynamics), we can see that the median shape of the BPM is similar to the median of the within-host ODE model (comparing Fig. 6(a) to Fig. 5(a)). However, we do see a secondary peak in the viral count in many of the infections. These correspond to a delayed, and high, peak in the lower respiratory tract. We also see that individuals with this second peak are those who survived infection. Indeed, while this second peak is higher, the infection is cleared earlier, meaning these individuals have a lower sustained viral load (and thus the area under the curve in the mortality proxy is lower).

For the posterior distribution of replication rates (*r*(*t*)) trajectories, in the majority of realisations *r*(*t*) between days zero and four was essentially constant (Fig. 6(b)), corresponding to the exponential growth of *v* (Fig. 6(a)). All of the trajectories also display a second increase in replication rates around day seven. In particular, individuals who died as a result of infection saw a slightly delayed and higher second peak in *r*(*t*), corresponding to the more sustained viral load exhibited (Fig. 6(a)). Studying the relationship between viral lifespan and peak replication rate (*r*(*t*)), there was minimal correlation between the two variables (Fig. 6(c)).

Study of the temporal dynamics for the uninfected target cells in the URT showed that most of the target cells are killed by virions, with the number of uninfected target cells dropping by two orders of magnitude. In the LRT, the number of target cells still decreases, but not to the extent as seen in the URT (Fig. S1(a)). The respective URT and LRT uninfected target cell trajectories paired with the observed free virion’s viral titre trajectories (Fig. S1(b)); viral trajectories peak higher and earlier in the URT, at around 10^7^TCID_50_/ml at approximately four days post infection, whereas peaks in viral trajectories for the LRT of 10^6^TCID_50_/ml, occurred later (7 to 8 days post infection). As we would expect, the infection peaks earlier and higher in the URT than the LRT. Numerous factors contribute to this, including the fluid movement required for initial viral particles to move to the LRT, advection of viral particles from the LRT back to the URT, the immune response being solely in the LRT, the infection in URT more quickly becoming saturated with infected cells (due to the considerably lower number of target cells), a lower production rate in the LRT, and the infection starting in the URT. The death of the host is from sustained immune response, as all total viral load trajectories are fairly similar up until approximately 6 days post infection, it is only the small differences in LRT viral load towards the end of the simulations that determines fatality.

When instead considering a case fatality rate of 20%, with individuals on average surviving longer given a lower case fatality rate, the range of viral lifespan was slightly shifted; those who died generally had a lifespan between 11.5 and 14 days (Fig. 3(b)). This contrasted to the viral lifespan distribution obtained in our main analysis (using a case fatality rate of approximately 73%), with more individuals being overwhelmed and dying, or alternatively clearing the virus after around 12 days (Fig. 3(b)). Despite these changes to the viral lifespan distribution, the viral dynamics are largely unaffected as the viral trajectories are simulated before fitting to the case fatality rate (further details in Section S4). Thus, a change in the case fatality rate only impacts the duration of infection within the host.

### 3.3 Human-to-human transmission probabilities

With estimates for the viral lifespan distribution and effective replication number from the parameter posterior distribution, we used our BPM to investigate viral mutation dynamics. Recall that we performed two sets of BPM realisations: one set with one million BPM realisations (1000 copies of the 1000 sample sets in the posterior parameter distribution) that each had an initial viral load of 10^6^ virions; a second set with 1000 BPM realisations (one for each sample comprising our parameter posterior distribution) that each had an initial viral load of 10^6^ × 10^6^.

From the first set of one million BPM realisations (see Section 2.3) we calculated the proportion of mutant strains over time (Fig. 7(a)) and probability of having at least one virion with *x* mutations over time (Fig. 7(b)). We found that virions with the required number of mutations for human-to-human transmission (three or more) made up a very small proportion of the viral load - around five orders of magnitude less than the strain with the next smallest proportion.

**Fig. 7.**
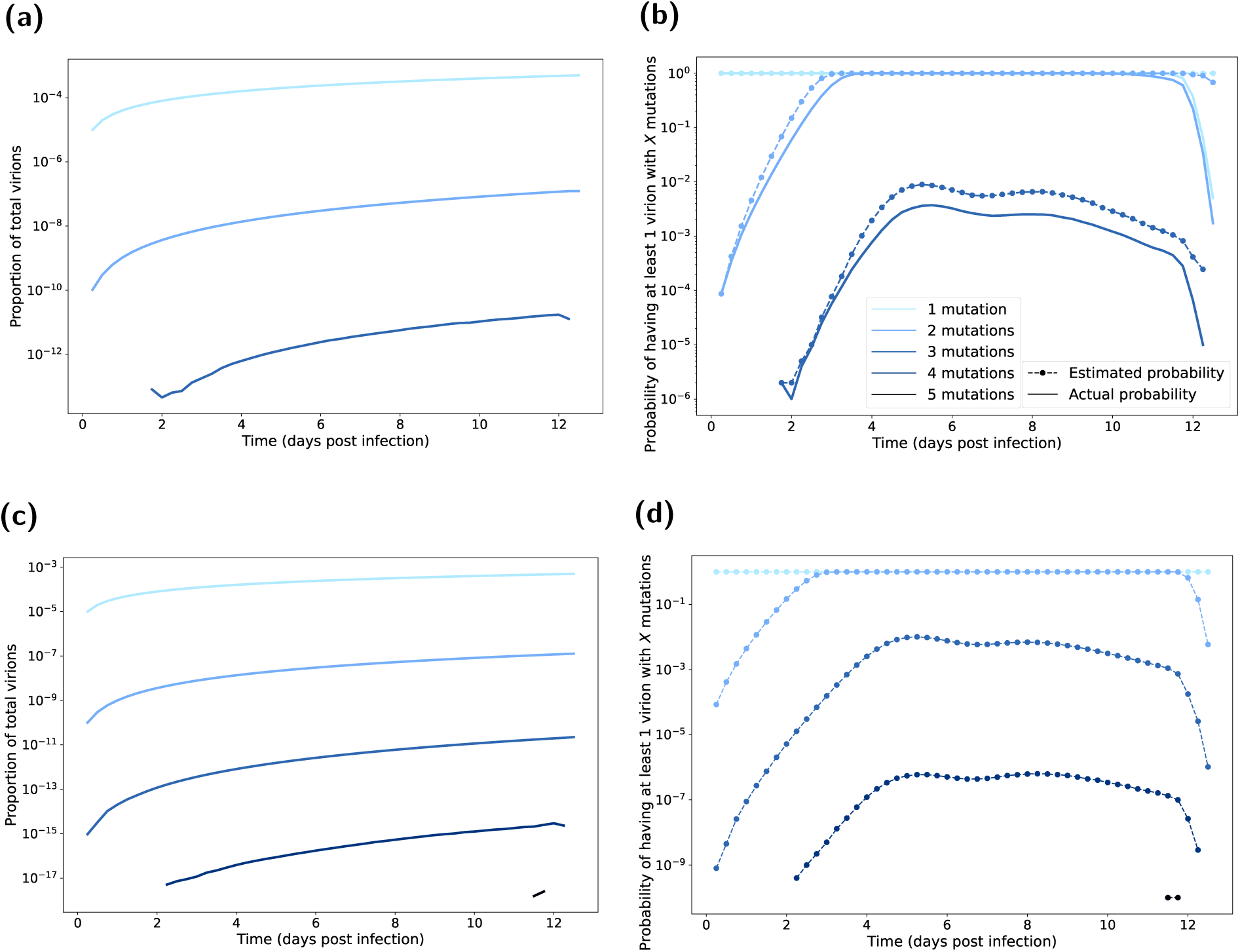
Mutated virion statistics with respect to time elapsed post infection, computed from BPM realisations. Line shading corresponds to the number of mutations (one mutation the lightest shading through to five mutations being the darkest shading). In all BPM simulations we fixed the probability of mutation at 10^−5^ per replication. In panels **(a&b)**, each realisation had an initial viral load of 10^6^. We ran 1000 realisations of each of the 1000 posterior parameter sets (Fig. 4). In panels **(c&d)**, each realisation had an initial viral load of 10^6^ × 10^6^. We ran one realisation of each of the 1000 posterior parameter sets (Fig. 4). **(a,c)** Proportion of total virions with the specified amount of mutations. There were a very small proportion of virions that have the required number of mutations to achieve droplet transmission (three or more mutations). **(b,d)** Probability of having a mutation strain. We present the estimated probabilities as the dashed lines with circle markers. We present the actual probabilities as solid lines. Probability estimate derivation follows that given in Section 2.2. The estimated probabilities are generally a clear upper bound on the true probabilities. Although not seen, we expect the only exceptions to be for higher number of mutations, where estimated probabilities for when these strains first occur could be initially below the actual probability; this reflected the dependence on the population of other mutants being more pronounced at lower numbers of virions (where the presence of a four mutation strain, for example, is almost purely from mutations from zero, one, two, three strain virions). Depending on the number of mutations in the initial infecting virions, there was a low probability of achieving the required number of mutations near the beginning on the infection lifespan (which would allow more replication of the mutant strains).

The derived probability approximation (Section 2.3) gave a generally sound upper bound, following a similar shape as the exact probability. Although unseen in these figures, other stochastic results indicate possible exceptions when a mutation first occurred and when the virus population died off near the end of the infection. This reflected the dependence on the population of other mutants being more pronounced at lower numbers of virions (where the presence of a four mutation strain, for example, is almost purely from mutations from zero, one, two, three strain virions). Once a strain becomes established within the population, the estimate as an upper bound is more robust as the majority of each strain comes from the replication of said strain (and not via mutation).

The set of BPM realisations with a higher starting load (of 10^6^ × 10^6^ initial virions) allowed for a more precise computation of the proportion of mutant virions (Fig. 7(c)). Although some virions mutated further along the pathway to droplet transmission (compared to the realisations with a lower initial viral load), they only made up a very small proportion of the total virion count. Similarly, for the estimates of the probability of observing at least one mutant with the required number of mutations (Fig. 7(d)), probabilities of obtaining either four or five required mutations for droplet transmission were very low across the entire infection duration.

In the case where we considered a lower case fatality rate of 20%, we observed similar mutation probabilities (Fig. S5). As expected, by lowering the case fatality rate the overall viral dynamics remained near identical for the majority of the infectious period. As such, the mutation probabilities were not affected until the end of the infection, by which point the peak of the mutation probabilities had already occurred.

## 4 Discussion

This paper presents a novel two-patch within-host model for an H5N1 influenza infection in a human host. Compared to existing literature, we explicitly model the lower and upper human respiratory tracts; this formulation enables us to mechanistically model the different biological responses to the infection in each tract. We also extend the earlier work of Russell *et al.* [5] to allow for more realistic modelling of virus mutations within a host. With these modelling developments we explored the risk of developing a human-to-human transmissible form of H5N1 influenza. Together these methods provide a general framework for combining within-host infection and within-host mutation models, which may be readily adapted to other (primarily respiratory) contagions – the adaptability of within-host models of this type to different pathogens has been showcased by Korosec *et al.* [37] , who repurposed the stochastic transmission-bottleneck model by Sigal *et al.* [16] (that had been applied to influenza A) to describe the survival probability of *de novo* SARS-CoV-2 mutations as a function of bottleneck size and selection coefficient.

The fitted within-host model displayed a preference for H5N1 influenza to spread in the LRT compared to the URT. This finding conforms with biological observations of a greater ease of spread for H5N1 influenza in the LRT (compared to the URT) [6, 7]. Also evident was the multi-modal nature of URT parameter posteriors. This is likely due to the URT behaving like a target-cell-limited model, in that the spread is only limited by the population of target-cells (as all of them become infected). Contagion dynamics are therefore less sensitive to the parameter values in the URT, resulting in the multi-modality of the parameter posterior distributions. Due to the higher target-cell numbers in the LRT, once the virus reaches the LRT the dynamics are much more sensitive to these parameters (*β_L_, p_L_*). As a consequence, the posterior has a much tighter peak around the mode. As previously stated, the qualitative shape of the median posterior predictive trajectory for viral titres in the URT is very similar to other models found in the literature [6, 13, 18, 36].

The analysis of the relationship between maximum effective replication number/growth rate, and peak viral load and infection lifespan revealed no correlation between these two variables. For the majority of infections, there was a second peak at around day eight corresponding to a delayed peak in the lower respiratory tract, which previous studies have indicated are to be biologically expected [20]. Under our default modelling assumptions all posterior predictive viral lifespans were less than 13 days. This is in agreement with the scenarios presented in Russell *et al.* [5], where they take the length of infection to be 10 days.

The modifications we made to an existing BPM for viral mutation, namely incorporating time-dependent replication rates and a realistic infectious duration distribution, gave comparable results to Russell *et al.* [5]. As the upper bound on the probabilities (of having at least one virion with *x* mutations) were of extremely low orders of magnitude, it seems highly unlikely that a typical human infection would lead to the arrival of a strain with four or five mutations. There is a much higher probability of having at least one virion with the (minimal) required three mutations, which may indicate that, with a large enough outbreak, we would expect a human-transmissible strain to evolve within at least one individual. Nonetheless, the proportion of virions with this strain is still expected to be very low and so transmission of such strains (even if present) is unlikely [5]. In contrast, strains with one or two mutations were generally highly prevalent amongst the virion population. Outbreaks in mammals (in particular the large number of infections in the dairy industry in the United States [38]), whose respiratory tracts are more similar to humans than avian species, may mean that human secondary infections from these animal cases are caused by a strain that is further along the mutation pathway to droplet transmissibility. Thus, there may be a higher than modelled risk of reaching the required number of mutations if a human is infected by a strain transmitted from other mammals, rather than birds. Russell *et al.* [5] considers differing initial mutations and also differing fitnesses of mutant strains. They conclude that although this does increase the proportions and probabilities stated, they are still sufficiently small such that these changes are unlikely to lead to a meaningful increase in the probability of human-to-human transmission, with which we concur.

Our work has not considered the probability of virions in the respiratory tracts being present in exhaled droplets and instead focused on the probability of mutating a droplet-transmissible variant. Consequently, the probabilities present in this paper are not equivalent to the probability of any H5N1 influenza infection in a human leading to a droplet transmissible virus. Nonetheless, our work does provide a framework for making this calculation. In the future, the development of a proxy or a further calculation from the results presented is required to make a conclusion on this transmission probability. In principle, any time that *p*^(^*^i^*^)^ *>* 0, there is a chance of human-to-human droplet transmission, with higher proportions of mutant strains corresponding to a higher likelihood of droplet transmission, though the exact relationship between these two entities is unclear. Our results show that it is unlikely, albeit not impossible, that a human infection of H5N1 influenza could lead to onwards transmission of a droplet transmissible strain. The probability results indicate that the presence of previous mutations at infection onset are more worrying than the development of the strain through mutations, as this would provide more time for a droplet transmissible strain to reach persistence levels in a host. Droplet transmissible strains mutating earlier in the infection pose a more significant threat as early mutations lead to higher proportions of mutant strains within the individual throughout the length of infection. Furthermore, an early mutation is likely to correspond with a longer period in which an infected individual is symptomatic with said mutant strain, and this leads to a higher probability of this mutant strain being droplet transmitted to another human.

The model we have presented is necessarily a simplified representation of reality. It is important that we consider the modelling assumptions made and their potential limitations. Here we elaborate on the implications of: the quality of the dataset used, estimation of the infection fatality ratio, estimation of the infection duration and initial viral load assumptions.

We note that there are limitations to the de Jong *et al.* [17] viral load dataset. Although our two-patch posterior predictions are very similar to other fitted models [6, 13, 18, 36], all within-host models for H5N1 influenza spread in human hosts that use this dataset suffer from a lack of parameter identifiability and biological certainty. In particular, due to the small size of the dataset, and because the majority of the patients died (even when given neuraminidase inhibitors), the average viral load may be much lower and viral lifespan much longer than is shown in our model. That being said, to provide an initial basis for the exploration of the effects of our novel two-tract within-host infection model, at the original time we conducted our study (mid-2024) we judged the de Jong *et al.* [17] H5N1 human influenza case viral load dataset to be appropriate data to use given the number of viral load samples it contained from a hospital patient cohort. We reaffirm the applicability of the developed methodology to any new viral load data generated from more recent human cases who contracted H5N1 avian influenza. An outbreak situation where datasets may be produced with viral load data sampled from multiple individuals who have unfortunately contracted HPAI H5N1 is the spillover case from dairy cattle in the United States; from April 2024, the Centers for Disease Control and Prevention (CDC) began reporting sporadic human cases of HPAI H5N1 in people who had exposure to infected dairy cows [38].

With regards to the estimation of the infection fatality ratio, at the time of writing, recorded cases are primarily hospitalisations and are therefore more likely to result in fatalities than unrecorded infections. Indeed, individuals could have been infected with H5N1 influenza and exhibited seasonal flu-like symptoms, or been asymptomatic. More recent studies also estimate a larger seroprevalence of H5N1 influenza in humans than previously calculated, implying that the actual fatality rate of an H5N1 influenza infection is lower than previously thought [39–41]. We assumed a default value for the infection fatality ratio that matched the mortality rate from the hospital patient cohort in the de Jong *et al.* [17] dataset (approximately 73%; 13 of the 18 patients). We acknowledge this H5N1 influenza mortality rate estimate for humans is likely an overestimate as these data corresponded to infection cases with severe illness. Nonetheless, our sensitivity analysis with a lower infection fatality ratio gave similar qualitative conclusions. Further infection data for H5N1 influenza viral titres in humans would be required for more accurate modelling estimates and conclusions. It is important that new cases are thoroughly documented, such that future H5N1 influenza models have improved accuracy, especially at the beginning and end of the infection dynamics.

Moreover, to link the infection fatality ratio to the in-host model we had to assume a relationship between fatality and observed viral dynamics. We took the cumulative viral load across both the URT and LRT as an appropriate proxy to represent mortality from cytokine storms and related immune responses. However, as there is no exact representation of mortality in the in-host model, we could also have justified using other proxies. For example, as the immune response is mainly caused from viral load in the LRT we could have instead weighted it higher in the integral. We anticipate that this would lead to extended lifespan distributions due to the delayed takeoff in the LRT. Nonetheless, we observed that the LRT peaks (the second peaks) already differentiate between surviving and dying patients and so our proxy already seems to be primarily influenced by LRT dynamics (Fig. 6). Further, extended lifespan distributions would only impact the proportion of virions with each mutation and not the total probability curves.

The fourth form of limitation relates to how pharmaceutical measures could impact the infectious duration of those infected with H5N1 influenza. Treatments, such as antivirals and neuraminidase inhibitors can reduce the viral load in individuals infected with H5N1 influenza exist and have been shown to be effective [18, 42, 43]. If infection with H5N1 influenza was caught early on then hospitalised individuals could be treated, with the resulting lower mortality rates and longer duration of infection (relative to the length of infection episodes that result in death) plausibly leading to higher than estimated probabilities of obtaining a droplet transmissible strain (similar treatments for COVID-19 patients led to higher mutation chances [44]). We note however, that in the dataset used, all individuals who presented with H5N1 influenza were subsequently given neuraminidase inhibitors, and yet all died due to their infection. Thus, it may be that in the majority of individuals, such treatments do not produce any meaningful increase in duration of infection of H5N1 influenza in humans.

Lastly, we had to make an assumption about initial viral load (which we fixed as 1.3×10^3^ TCID_50_/ml). Given the infection data used is primarily centred around the peak of infection, our inference is most strongly determined by this peak behaviour. As a consequence, the early growth rate corresponds to parameter estimates that give the ‘correct’ peak height and time for the data, given the assumed initial viral load. A change in the initial viral load would change the growth rate with a negative correlation to the initial viral load. Nonetheless, we anticipate the viral lifespan distribution would be similar for different assumed initial viral load values (as it is a function of the peak time and area under the curve, which should not be affected much by the early rates of growth).

For the mutations model, a change in the initial viral load would result in the same proportions as shown in our results (as they are primarily dependent on the mutation probability). However, the curves for mutant strain *probabilities* would be shifted up and towards the left such that there are increased probabilities of observing strains with higher numbers of mutations earlier in the infection. Furthermore, the mutation probability estimates are based on the simulated dataset of posterior predictive trajectories. We observe in Fig. 5 that there are orders of magnitude more variation in the viral titre dataset. We anticipate that including this uncertainty would have similar impacts to a change in the initial conditions – trajectories including the lower viral titres would lead to mutation probability curves shifted down and towards the right and trajectories including the upper viral titres would shift the curves up and towards the left.

In addition to the aforementioned ideas for additional work, another direction for further investigations is the application of the model framework to infectious episodes in immunocompromised individuals. During the COVID-19 epidemic, immunocompromised individuals were a large cause for concern for the creation of new variants due to their longer duration of infection [44–46]. To our knowledge, there have been no reported cases of an immunocompromised individual being infected by H5N1 influenza, and thus it is unclear how they would respond to the infection. As previously stated, the main cause of death in those who contracted H5N1 influenza is currently believed to be cytokine storm. This was also the leading cause of death from the Spanish flu epidemic in 1918-1920, but the fatality rate was lower for the immunocompromised as they did not exhibit a sufficient immune response to cause a cytokine storm [47, 48]. As a result, it may be that immunocompromised individuals are able to sustain longer periods of infection, thus giving a larger probability of a human-to-human transmissible strain mutating during their infection period. It is also possible that the virus simply overwhelms the body of the immunocompromised, leading to rapid death, and thus little chance of producing mutant strains of H5N1 influenza. Our current datasets are unable to distinguish between these possible outcomes. The literature also shows that infections from H5N1 influenza can spread to other organs and parts of the body [17]; it is likely that more detailed mortality modelling would need to take this into account with different mortality modelling methods for immunocompromised people.

In this paper we have provided a model framework that gives the basis for the calculation of the probability that the increased prevalence of influenza A(H5N1) in both birds and mammals leads to a human infection that develops the ability for droplet transmission. These advancements in modelling tools can help us determine how pandemic preparedness resources may be best focused between infection directly from avian hosts or from mammalian hosts. Indeed, our process is not just relevant to H5N1 influenza, but also for any pathogen for which within-host mutations are a concern.

## Data Availability

All data utilised in this study are publicly available, with relevant references and data repositories provided. The code repository for the study is available at: https://github.com/joshlooks/avianflu.

## Author contributions

**Daniel Higgins:** Data curation, Formal analysis, Methodology, Software, Validation, Visualisation, Writing - Original Draft, Writing - Review & Editing.

**Josh Looker:** Data curation, Formal analysis, Methodology, Software, Validation, Visualisation, Writing - Original Draft, Writing - Review & Editing.

**Robert Sunnucks:** Data curation, Formal analysis, Methodology, Software, Validation, Visualisation, Writing - Original Draft, Writing - Review & Editing.

**Jonathan Carruthers:** Conceptualisation, Methodology, Supervision, Visualisation, Writing - Review & Editing.

**Thomas Finnie:** Conceptualisation, Methodology, Supervision, Visualisation, Writing - Review & Editing.

**Matt J. Keeling:** Conceptualisation, Methodology, Supervision, Visualisation, Writing - Review & Editing.

**Edward M. Hill:** Conceptualisation, Methodology, Supervision, Visualisation, Writing - Review & Editing.

## Data accessibility

All data utilised in this study are publicly available, with relevant references and data repositories provided.

## Code availability

The code repository for the study is available at: https://github.com/joshlooks/avianflu.

Archived code available at: https://doi.org/10.5281/zenodo.13385415.

## Supporting Information

### S1 Additional tables

**Table S1.**
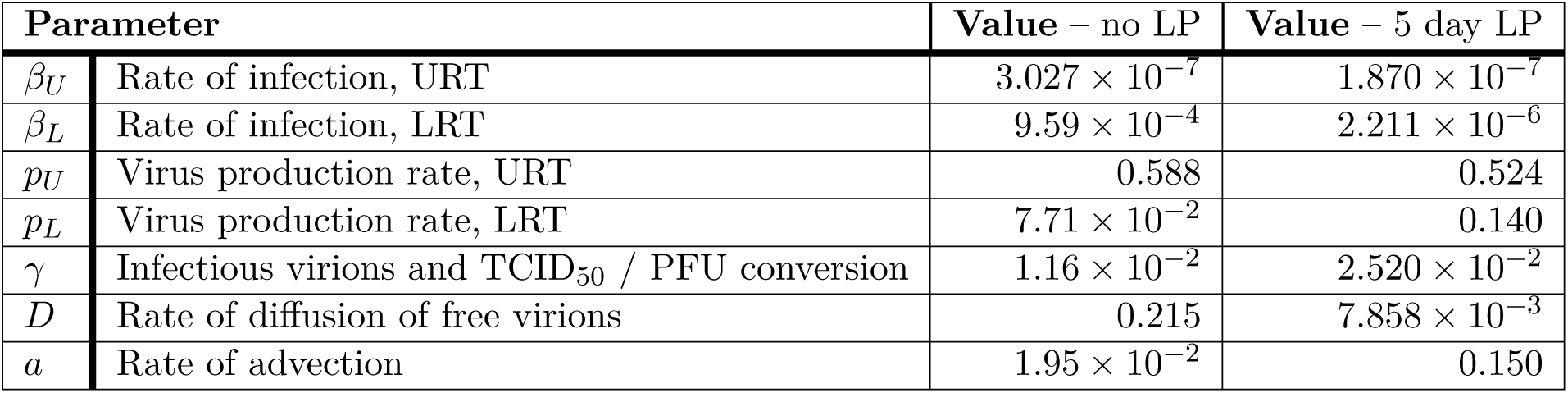
Least-squares fit parameters: and their descriptions. Parameters fitted to the data in the log-space using the ‘scipy.optimize’ library in Python. We present values under two different assumptions for mapping days post-infection in the model to the de Jong *et al.* [1] viral load data that is dependent on ‘days since onset of illness’. We amend the de Jong *et al.* [1] viral load data to be in terms of days post-infection rather than ‘days since onset of illness’ by applying either no latent period or a 5 day latent period (LP).

### S2 Additional figures

**Fig. S1.**
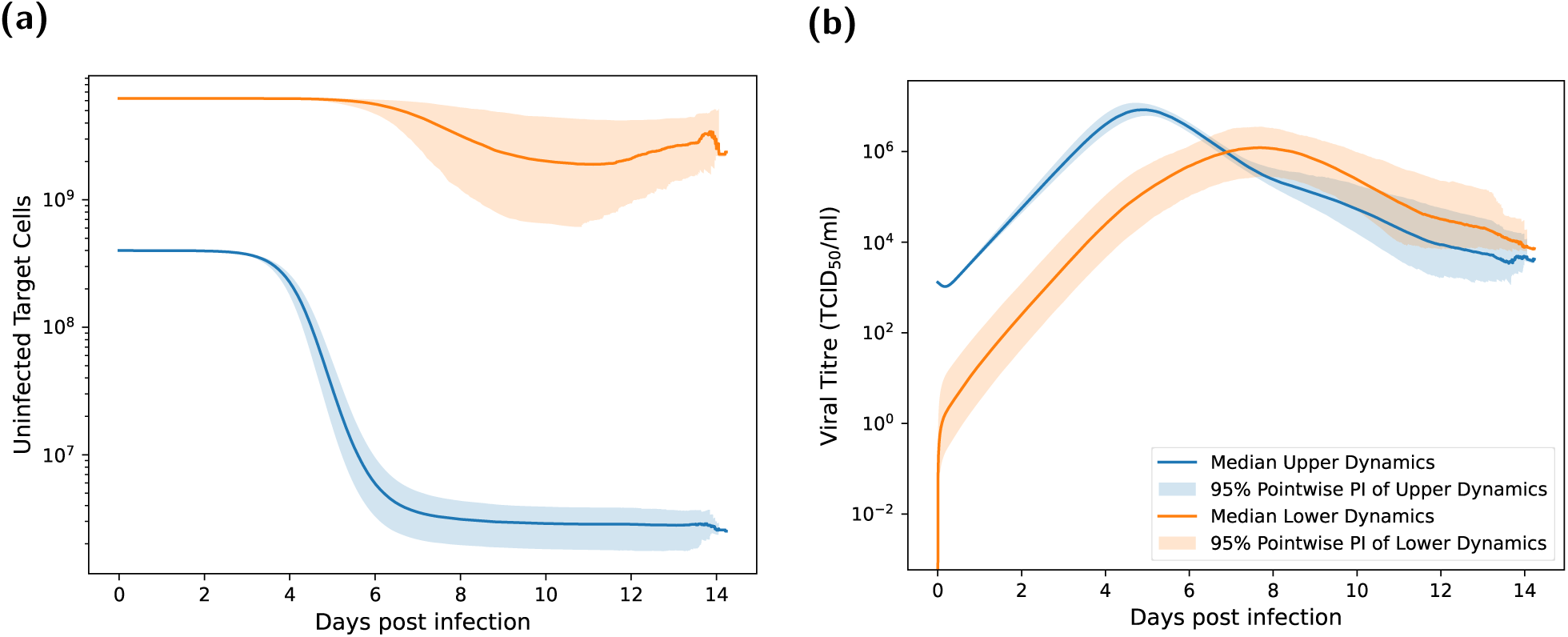
Compartmental trajectories for uninfected target cells and free virions. Both plots show the median and 95% pointwise predictive interval of the 1000 posterior trajectories, with the blue lines representing dynamics of the URT and the orange lines representing dynamics of the LRT. **(a)** The target cell count trajectories. Most of the target cells in the URT are killed by virions, with the number of uninfected target cells dropping by two orders of magnitude. In the LRT, the number of target cells still decreases, but not to the extent as seen in the URT. **(b)** The free virion’s viral titre trajectories. Viral trajectories peak higher and earlier in the URT, at around 10^7^TCID_50_/ml at approximately four days post infection. Peaks in viral trajectories for the LRT are lower and occurred later, at roughly 10^6^TCID_50_/ml after 7-8 days post infection.

#### S3 Robustness of model outcomes when applying an incubation period to viral load data

Here we present results assuming an incubation period of five days when calibrating model outputs in terms of days post-infection to the de Jong *et al.* [1] viral load data that was dependent on ‘days since onset of illness’. An incubation period of five days equals the median incubation period reported by a retrospective cohort study of 18 influenza virus (H5N1) cases in China [2]. Compared to our main analysis findings, under this alternative incubation period assumption we observed a similar posterior distribution for *β_U_ , p_U_* and *γ* (Fig. S2). Although the *β_L_* has most of its distribution mass much higher than *β_U_* , and *p_L_* lower than *p_U_* , we note that the differences in order of magnitude between them have decreased. The rate of diffusion is now much smaller and the rate of advection much higher. These changes are likely due to the five day latent period requiring delayed and possibly smaller take-off in the lower respiratory tract than the results assuming a zero day incubation period.

For the posterior distribution of replication rates (*r*(*t*)) trajectories, in the majority of realisations *r*(*t*) between days zero and six post infection was essentially constant, corresponding to the exponential growth of *v* (Fig. S3). In contrast to the main analysis, there was not a second increase in replication rates. Instead, all replication rate trajectories had a consistent decline until about day 12 post infection, beyond which they plateaued (Fig. S3(b)).

Studying the relationship between viral lifespan and peak replication rate (*r*(*t*)), there was once a negative correlation between the viral lifespan and peak replication rate (Fig. S4).

Our general conclusions from the branching process model analysis also remained unchanged (Fig. S5).

**Fig. S2.**
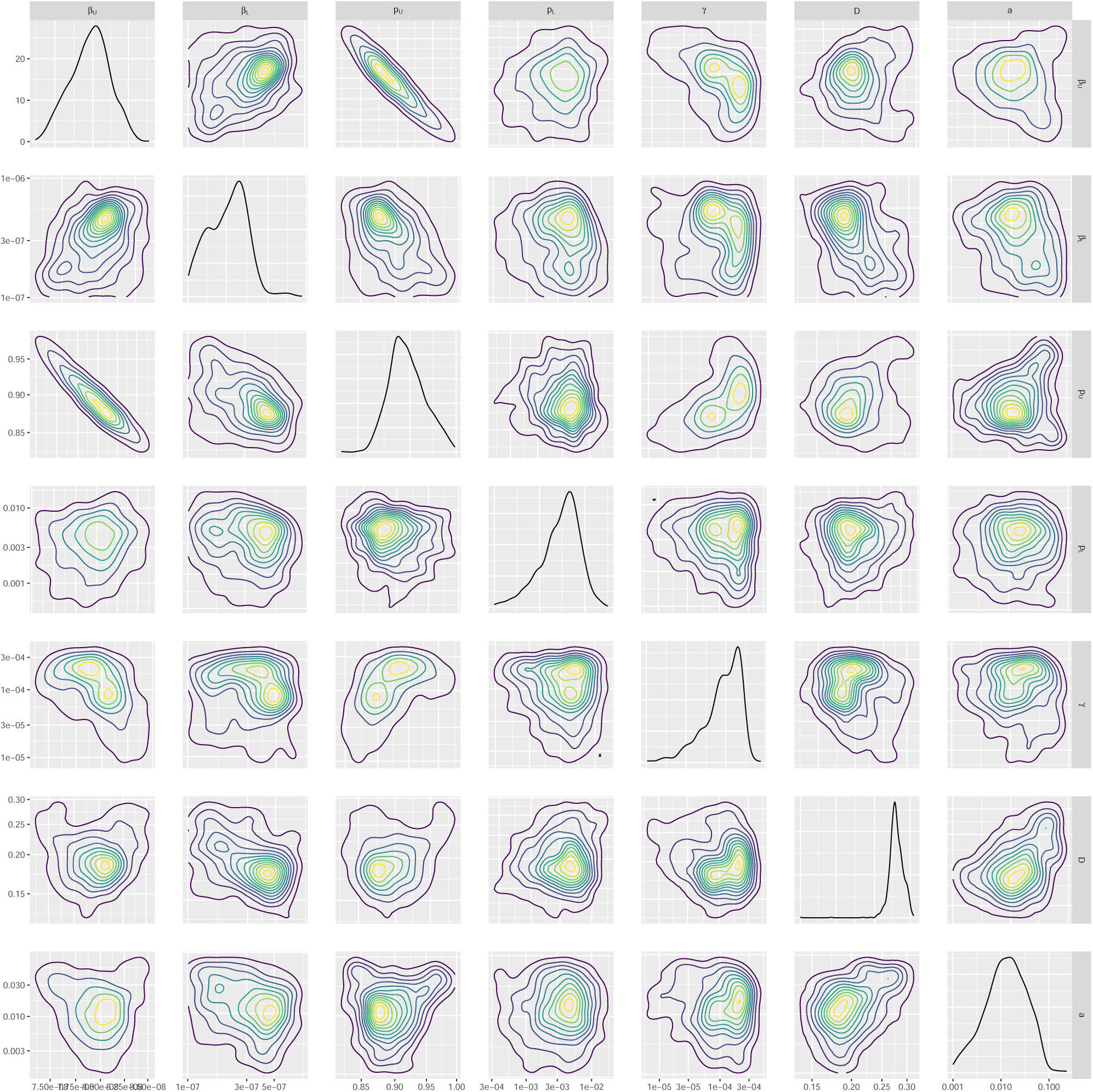
Parameter posterior distributions. We obtained 1000 samples of the target posterior distribution using the ABC-SMC-MNN method outlined in Section 2.1.5. Diagonal panels show the marginal distributions for: rate of infection in the URT (*β_U_*) and the LRT (*β_L_*), virus reproduction rate in the URT (*p_U_*) and the LRT (*p_L_*), viral uptake rate between infection and viral titre (*γ*), rate of diffusion of free virions (*D*) and the rate of advection (*a*), respectively. Off-diagonal panels show bi-parameter distributions, where the contour shading intensity corresponds to the probability density value (lighter for higher probability density). Parameters (*β_L_, p_L_*) in the LRT tended to be higher than the URT (*β_U_ , p_U_*), agreeing with the biological preference for H5N1 influenza to infect the LRT.

**Fig. S3.**
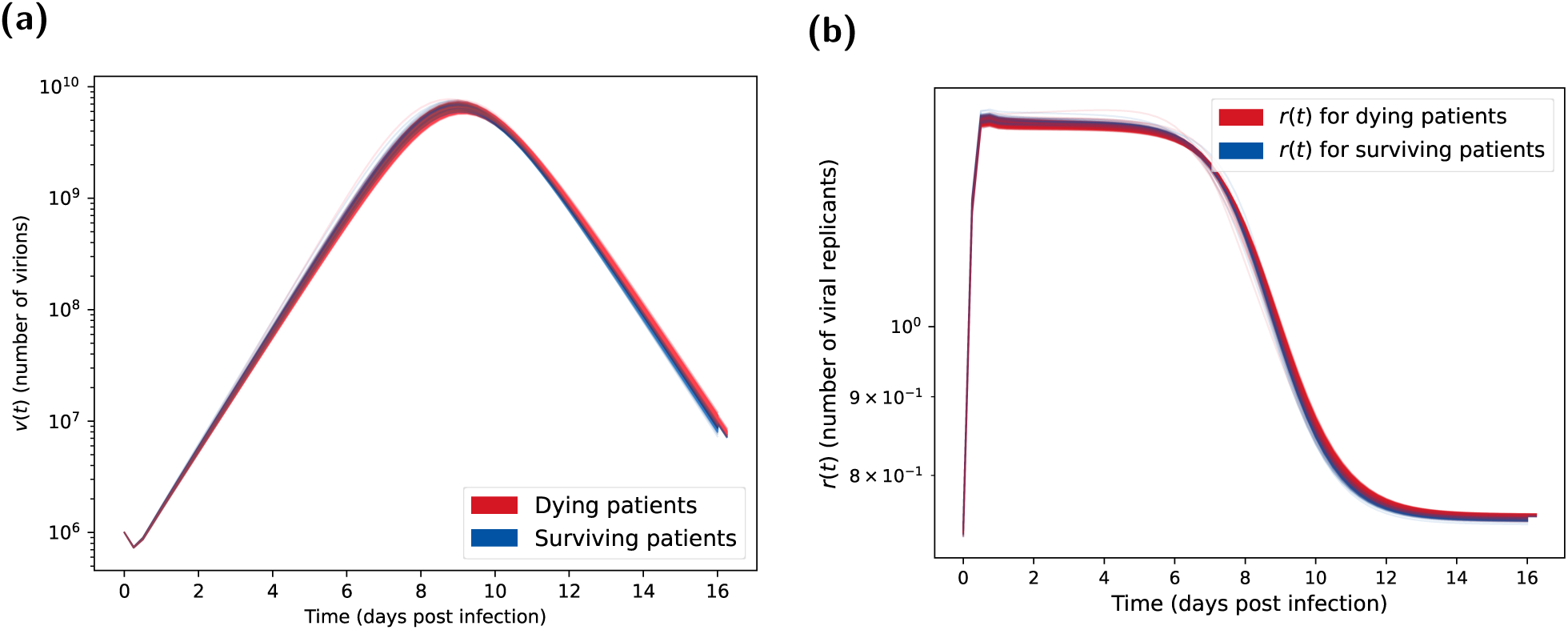
Posterior predictions for number of virions and number of viral replicants (*r*(*t*)) assuming a five day latent period. Both plots show the 1000 posterior trajectories, with the blue lines representing H5N1 influenza patients who survive the infection (cleared the virus) and the red lines representing patients who died due to the infection (where the distinction is made using the method in Section 2.1.6). **(a)** Virion count distribution found using the parameter posterior in Fig. 4. We see similar trends between surviving and deceased patients. We calculated these viral count trajectories from the one million BPM realisations each with an initial viral load of 10^6^ and a mutation chance of 10^−5^ per replication. **(b)** Distribution of *r*(*t*) calculated from the posterior predictive distribution shown in Fig. 5(a). Surviving patients tended to have slightly higher *r*(*t*) in the peak, which then declined below one (indicating a decreasing virion count) marginally faster than for dying patients.

**Fig. S4.**
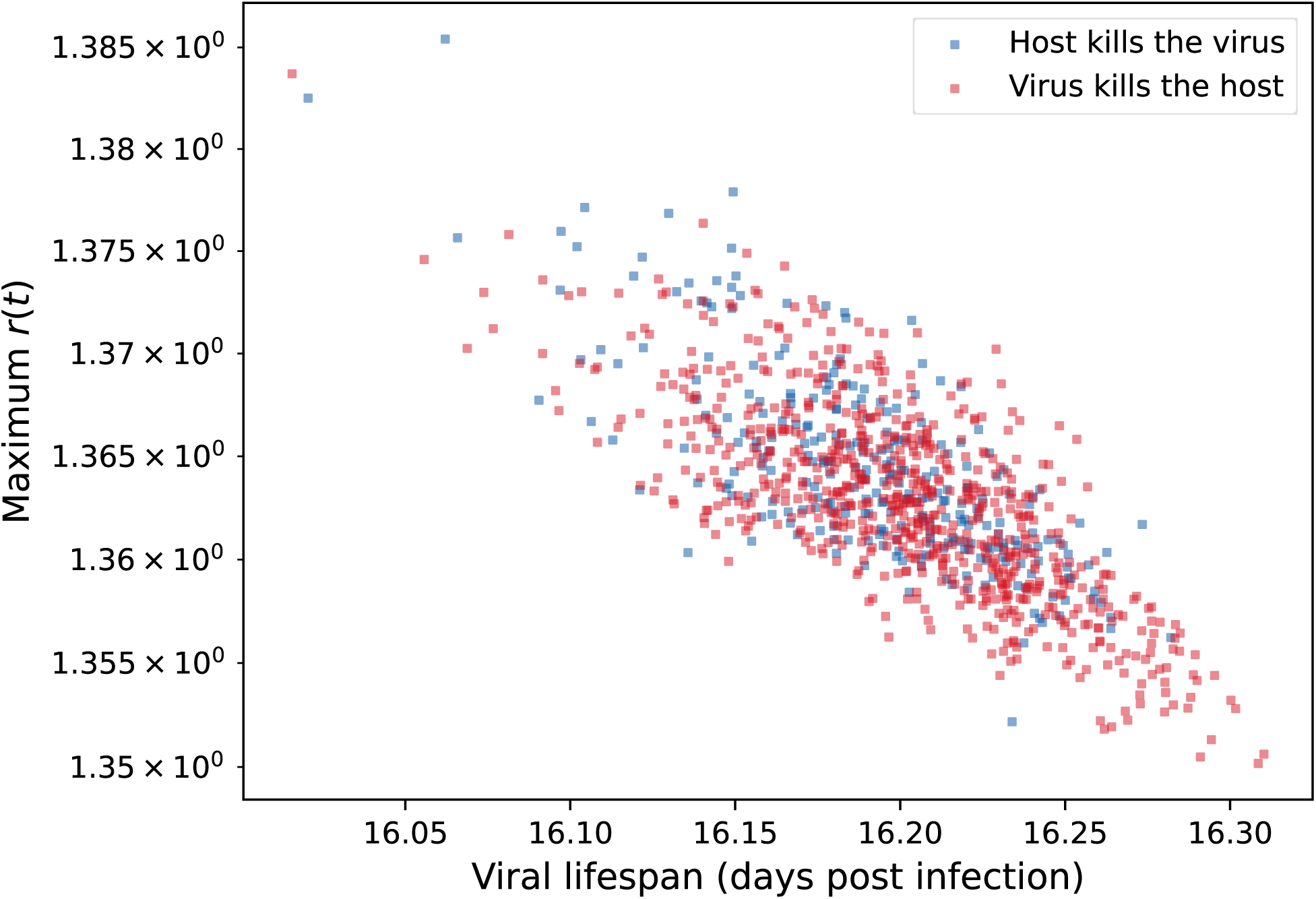
Maximum *r* value vs viral lifespan assuming a five day latent period. We observe a negative correlation between maximum *r* value and the viral lifespan. Blue circles represent H5N1 influenza patients who survive the infection (cleared the virus). Red circles represent patients who died due to the infection (where the distinction is made using the method in Section 2.1.6). Surviving individuals tended to have lower viral lifespans.

**Fig. S5.**
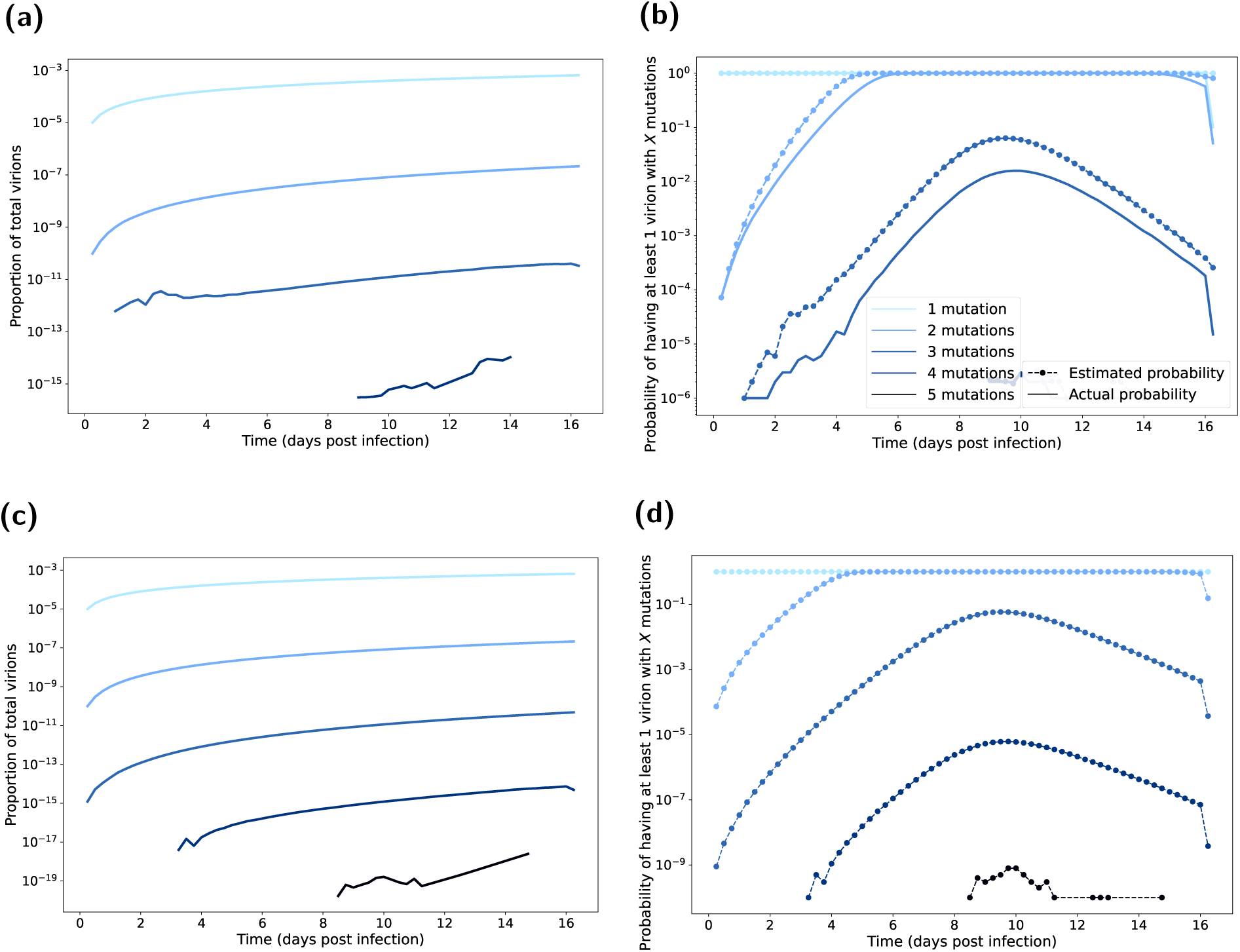
Mutated virion statistics with respect to time elapsed post infection, computed from BPM realisations assuming a five day latent period. Line shading corresponds to the number of mutations (one mutation the lightest shading through to five mutations being the darkest shading). In all BPM simulations we fixed the probability of mutation at 10^−5^ per replication. In panels **(a&b)**, each realisation had an initial viral load of 10^6^. We ran 1000 realisations of each of the 1000 posterior parameter sets (Fig. 4). In panels **(c&d)**, each realisation had an initial viral load of 10^6^ × 10^6^. We ran one realisation of each of the 1000 posterior parameter sets (Fig. 4). **(a,c)** Proportion of total virions with the specified amount of mutations. There were a very small proportion of virions that have the required number of mutations to achieve droplet transmission (three or more mutations). **(b,d)** Probability of having a mutation strain. We present the estimated probabilities as the dashed lines with circle markers. We present the actual probabilities as solid lines. Probability estimate derivation follows that given in Section 2.2. The estimated probabilities are a clear upper bound on the true probabilities. Depending on the number of mutations in the initial infecting virions, there was a low probability of achieving the required number of mutations near the beginning on the infection lifespan (which would allow more replication of the mutant strains).

#### S4 Robustness of model outcomes to a lower case fatality rate

Here we present the results when conducting the analysis assuming a lower case fatality rate of 20%. We took the case fatality rate estimate of 20% from Dobrovolny *et al.* [3] for individuals treated with neuraminidase inhibitors.

Inspecting the viral dynamics results, with the viral trajectories being simulated before fitting to the case fatality rate, we found the viral dynamic results assuming a lower case fatality rate to be similar to the main analysis (Figs. S6 and S7).

Comparing the estimated mutation probabilities between the main analysis and when assuming the lower case fatality rate of 20%, we observed similar mutation probabilities (Fig. S8). The overall viral dynamics remained near identical for the majority of the infectious period. Consequently, the mutation probabilities were not affected until the end of the infection, by which point the peak of the mutation probabilities had already occurred.

**Fig. S6.**
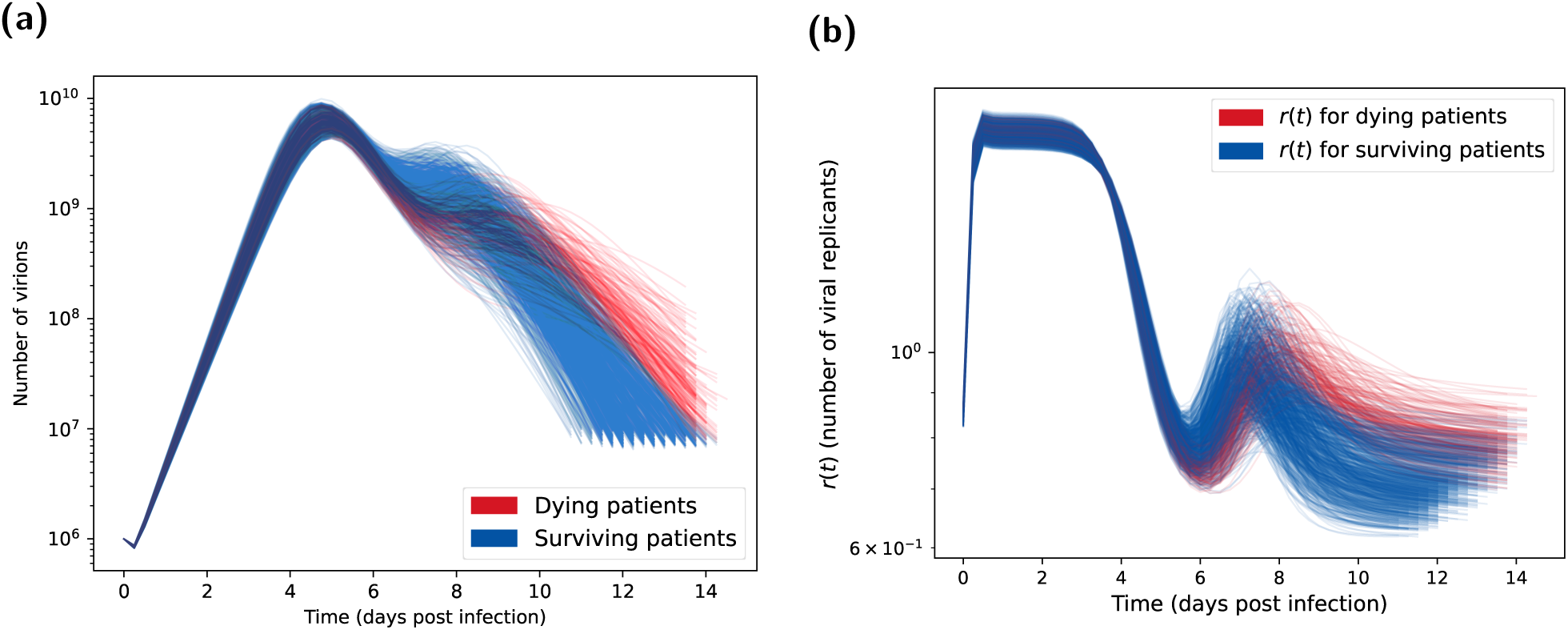
Posterior predictions for number of virions and number of viral replicants (*r*(*t*)) assuming a lower case mortality rate (20% instead of 73%). Both plots show the 1000 posterior trajectories, with the blue lines representing H5N1 influenza patients who survive the infection (cleared the virus) and the red lines representing patients who died due to the infection (where the distinction is made using the method in Section 2.1.6). **(a)** Virion count distribution found using the parameter posterior in Fig. 4. The viral count trajectories for deceased patients are lower and more sustained than surviving patients. We calculated these viral count trajectories from the one million BPM realisations each with an initial viral load of 10^6^ and a mutation chance of 10^−5^ per replication. **(b)** Distribution of *r*(*t*) calculated from the posterior predictive distribution shown in Fig. 5(a). Surviving patients tended to have higher *r*(*t*) in the second peak around day 7 on infection, which then declined below one (indicating a decreasing virion count) more rapidly than for dying patients.

**Fig. S7.**
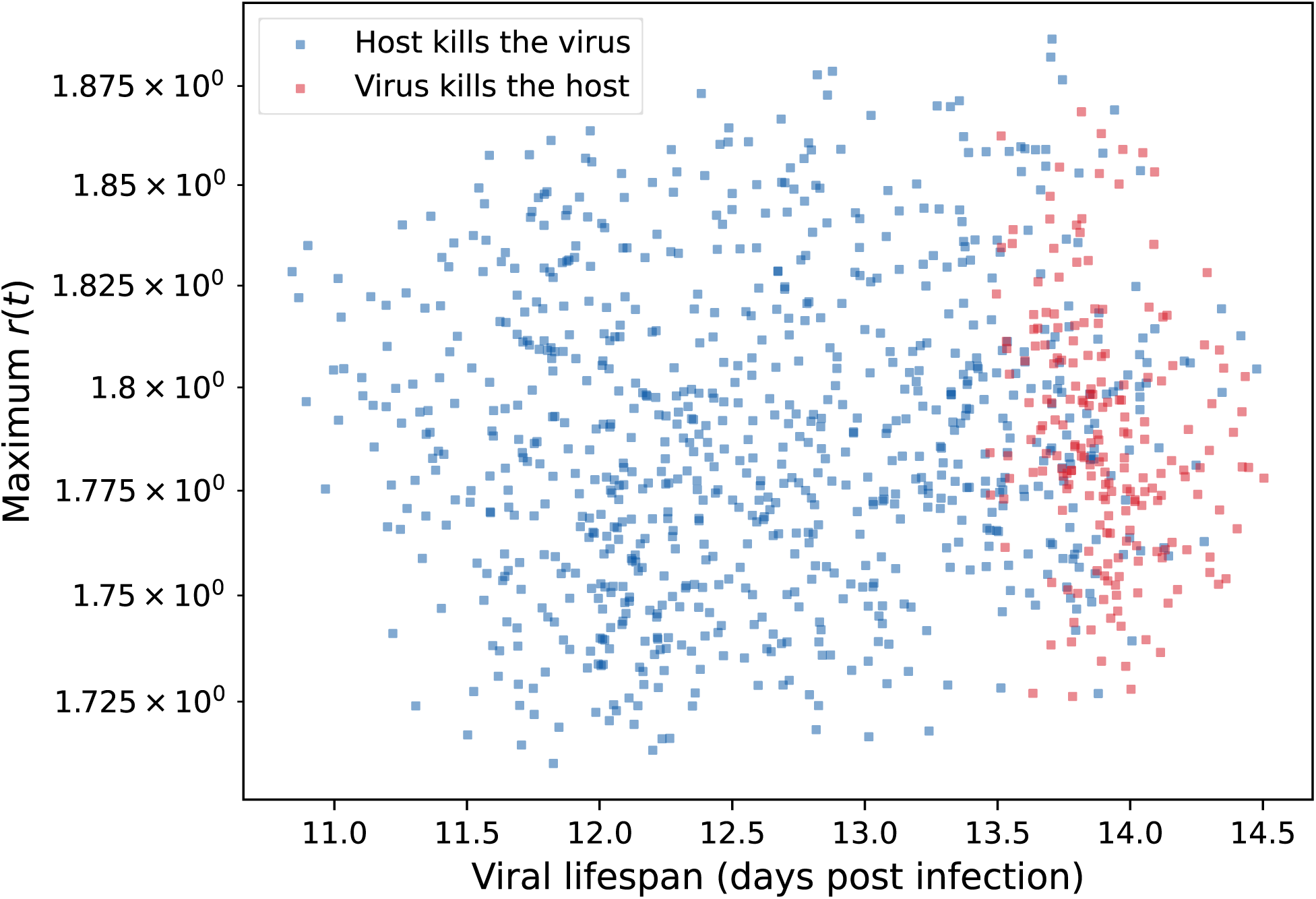
Maximum *r* value vs viral lifespan assuming a lower case mortality rate (20% instead of 53%). We found no strong correlation between maximum *r* value and the viral lifespan. Blue circles represent H5N1 influenza patients who survive the infection (cleared the virus). Red circles represent patients who died due to the infection (where the distinction is made using the method in Section 2.1.6). Surviving individuals tended to have lower viral lifespans.

**Fig. S8.**
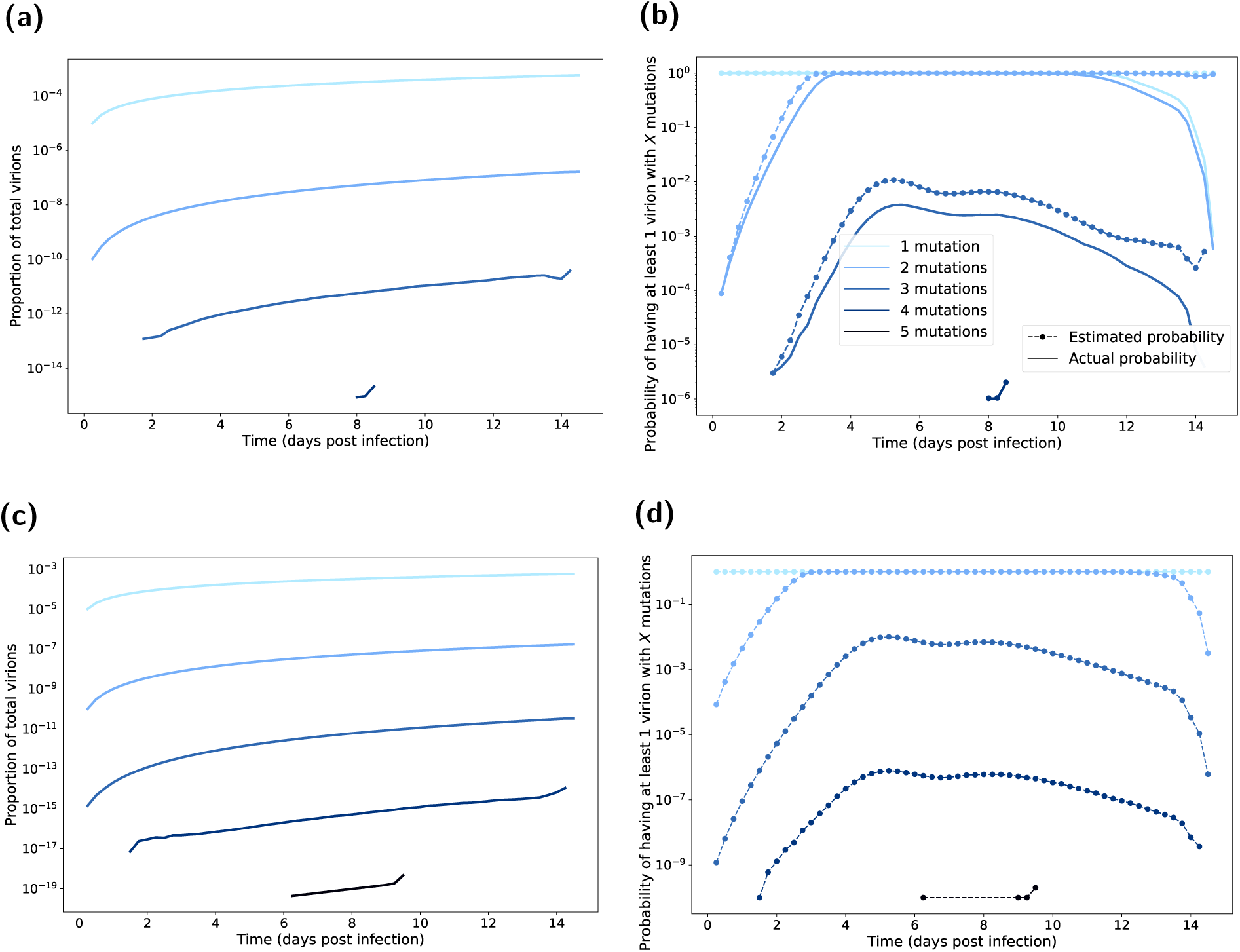
Mutated virion statistics with respect to time elapsed post infection, computed from BPM realisations assuming a lower case mortality rate (20% instead of 73%). Line shading corresponds to the number of mutations (one mutation the lightest shading through to five mutations being the darkest shading). In all BPM simulations we fixed the probability of mutation at 10^−5^ per replication. In panels **(a&b)**, each realisation had an initial viral load of 10^6^. We ran 1000 realisations of each of the 1000 posterior parameter sets (Fig. 4). In panels **(c&d)**, each realisation had an initial viral load of 10^6^ × 10^6^. We ran one realisation of each of the 1000 posterior parameter sets (Fig. 4). **(a,c)** Proportion of total virions with the specified amount of mutations. There were a very small proportion of virions that have the required number of mutations to achieve droplet transmission (three or more mutations). **(b,d)** Probability of having a mutation strain. We present the estimated probabilities as the dashed lines with circle markers. We present the actual probabilities as solid lines. Probability estimate derivation follows that given in Section 2.2. The estimated probabilities are a clear upper bound on the true probabilities. Depending on the number of mutations in the initial infecting virions, there was a low probability of achieving the required number of mutations near the beginning on the infection lifespan (which would allow more replication of the mutant strains).

