## Supporting Information for "Introducing a framework for within-host dynamics and mutations modelling of H5N1 influenza infection in humans"

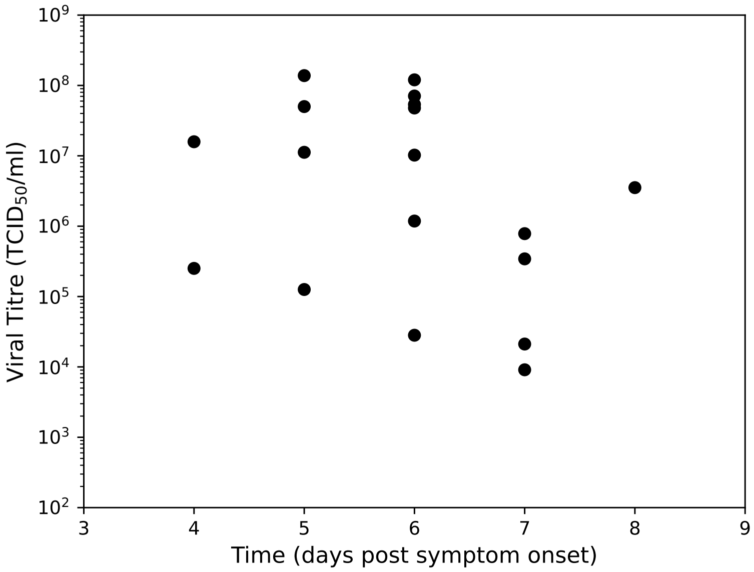


**Fig. 1. Viral titres from pharyngeal swabs of hospitalised H5N1 influenza patients in Vietnam.** Data from de Jong *et al.* [17], where viral RNA loads were measured in throat swabs obtained at admission from 18 H5N1 patients. We used this dataset to calibrate all models included in this paper.

$$\frac{dT_U}{dt} = -\beta_U T_U V_U$$
$$\frac{dE_U}{dt} = \beta_U T_U V_U - gE_U$$
$$\frac{dI_U}{dt} = gE_U - dI_U$$
$$\frac{dV_U}{dt} = p_U I_U - cV_U - \gamma\beta_U T_U V_U - D(V_U - V_L) + aV_L$$

$$\frac{dT_L}{dt} = -\beta_L T_L V_L$$
$$\frac{dE_L}{dt} = \beta_L T_L V_L - gE_L$$
$$\frac{dI_L}{dt} = gE_L - dI_L$$
$$\frac{dV_L}{dt} = p_L I_L - cV_L - \gamma\beta_L T_L V_L - kV_L X + D(V_U - V_L) - aV_L$$
$$\frac{dX}{dt} = fV_L + rX\omega X$$

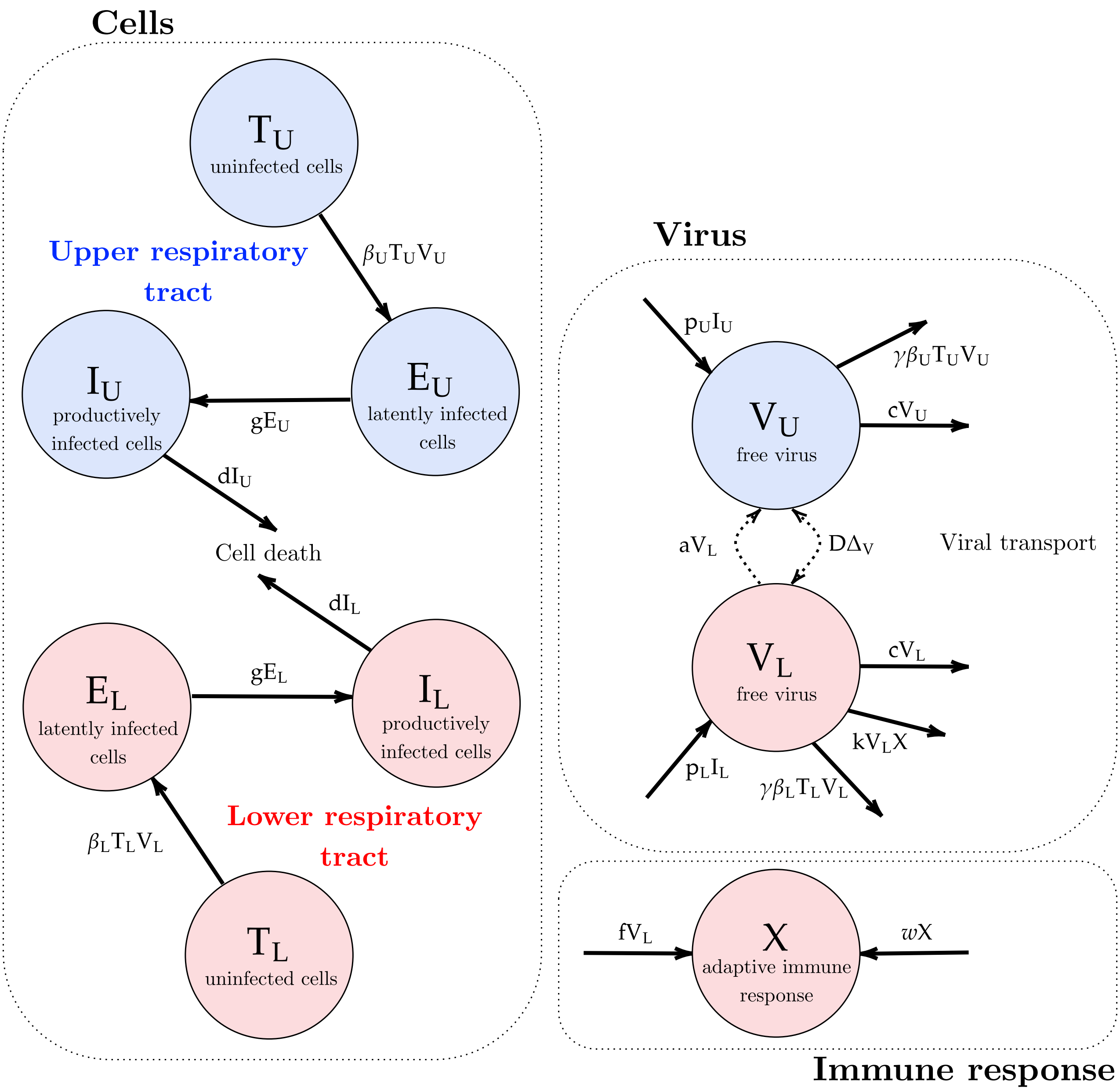


**Fig. 2. LRT and URT explicit within-host respiratory infection model schematic.** Compartments listed are uninfected/target cells ($T$), free virions ($V$), eclipse/latent cells ($E$), infected/virion-producing cells ($I$) and the adaptive immune response ($X$). Note that the subscripts $U, L$ represent the URT- and LRT-based compartments respectively. The different colours represent the processes in the URT (in blue) and in the LRT (red). Arrows show the spread of the contagion through the host. The dashed arrows in the virus compartment indicate the coupling of the two patches through advection and diffusion. Parameters descriptions are found in Table 1.

with $\beta_U$ and $\beta_L$ the rate of infection in the URT and LRT, $g$ the latent transition rate of infected cells, $d$ the mortality rate of infected virus producing cells , $p_U$ and $p_L$ the virus production rate in the URT and LRT, $c$ the morality rate of free virions, $\gamma$ the conversion rate between infection and viral titre (also referred to as the viral uptake rate), $f$ the recruitment rate of adaptive immune response, ~~$r$~~ $\omega$ the expansion rate of adaptive immune response and $k$ the kill rate of adaptive immune response, $D$ the rate of diffusion and $a$ the rate of advection.

##### 2.1.4 Model parameterisation from the literature

We obtained values from the literature for a subset of parameters in our ODE model. Specifically, we used the parameter point estimates reported by Dobrovolny *et al.* [18] to set the latent state duration of infected cells ($1/g$) as 1/4 days, the lifespan of infected virus producing cells ($1/d$) to be 1/5.2 days and the lifespan of free virions ($1/c$) as 1/2 day. We also highlight that Dobrovolny *et al.* [18] noted that their values were consistent with other research in the area.

For the immune parameters, we took the approach found in Handel *et al.* [20]. Although this was fitted to mice data, studies have shown that the mice immune system is a suitable analogue for the immune system found in humans *in vivo* [21]. Further, parameters are likely transferable through the comparison of mice and human metabolic rates - mice have a metabolic rate seven times that of humans [22]. We converted from plaque-forming unit (pfu) into $\text{TCID}_{50}$ (Tissue culture infectious dose 50%), with pfu being proportional to TCID50 by a factor of 0.56 [23].

It was also important to select an initial number of target cells and initial viral load. We took the estimated values of $T_U = 4 \times 10^8, T_L = 6.25 \times 10^9$ from Ciupe and Tuncer [12], which were calculated using the average surface area of an epithelial cell and of the human respiratory tract. We took the initial viral load ($V_0 = 1.3 \times 10^3$ $\text{TCID}_{50}$/ml) from the fitted values of the single-target-cell model in Dobrovolny *et al.* [6].

There was little information in the literature regarding rate of infection $\beta_U$ and $\beta_L$, virus production rate $p_U$ and $p_L$, conversion rate (rate of viral uptake) between infection and viral titre $\gamma$, rate of diffusion $D$ or rate of advection $a$. These parameters of interest were also chosen as they have been found to have the biggest impact on the observed disease dynamics [6, 13, 18]. It is also worth noting that setting $\gamma = 0$ leads to similar results (and is normally ignored in human models [13, 18, 20]). This parameter represents the uptake rate between the viral titre (in $\text{TCID}_{50}$) and the number of free virions used to infect a target-cell. Setting this parameter to zero indicates that there is no noticeable change in the viral titre due to the infection of target-cells. By re-introducing this parameter (allowing it to be non-zero), we gained an extra degree of freedom in the model that allowed for more biologically realistic parameter values and peak shapes to be observed during parameter fitting.

We state the default model parameters, for non-fitted parameters, in Table 1. Some of the selected parameter values are similar to literature values for models fitted to H1N1 infection data within humans [24–26]. However, previous studies on H5N1 influenza infection in humans found that these values gave good fits to the data, and that the other aforementioned parameters that we fitted for were the main contributors to viral dynamics [6, 18].

##### 2.1.5 Model calibration and parameter inference

To calibrate the model, we made use of the dataset outlined in Section 2.1.1. We note that these data correspond to the viral load in the URT only, meaning we could only fit the model dynamics based on the URT compartments. Parameter identifiability is a problem for most mathematical biology models, and this was especially true for our fitting process as we had less than 20 data points available, all of

**Table 1. List of model parameters and their descriptions.** For fixed parameters we state their value and associated references. For inferred parameters we list their prior distribution (we use $lU$ as a notation for the log-uniform distribution). We provide unit information for each parameter in parenthesis after the parameter description.

| **Parameter** | | Value | Prior |
|---|---|---|---|
| $\beta_U$ | Rate of infection, URT ($\text{day}^{-1}$) | - | $l\text{U}(1\times10^{-8}, 1\times10^{-6})$ |
| $\beta_L$ | Rate of infection, LRT ($\text{day}^{-1}$) | - | $l\text{U}(1\times10^{-7}, 1\times10^{-5})$ |
| $1/g$ | Productively infected cells (days) | 1/4 [18] | - |
| $1/d$ | Lifespan of infected, virus-producing cells (days) | 1/5.2 [18] | - |
| $p_U$ | Virus production rate, URT ($\text{day}^{-1}$) | - | $l\text{U}(1\times10^{-4}, 1)$ |
| $p_L$ | Virus production rate, LRT ($\text{day}^{-1}$) | - | $l\text{U}(1\times10^{-4}, 1)$ |
| $1/c$ | Lifespan of free virions (days) | 1/2 [18] | - |
| $\gamma$ | Conversion between infectious virions and $\text{TCID}_{50}$ / PFU (unitless) | - | $lU(1\times10^{-6}, 2\times10^{-3})$ |
| $f$ | Recruitment rate of adaptive immune response ($\text{day}^{-1}$) | $0.56\times2.8\times10^{-6}/7$ [20] (fig 6) | - |
| $r$ | Expansion rate of adaptive immune response ($\text{day}^{-1}$) | 0.27/7 [20] (fig 6) | - |
| $k$ | Kill rate of adaptive immune response ($\text{day}^{-1}$) | 20 [20] | - |
| $D$ | Rate of diffusion of free virions ($\text{day}^{-1}$) | - | $l\text{U}(1\times10^{-3}, 1)$ |
| $a$ | Rate of advection ($\text{day}^{-1}$) | - | $l\text{U}(1\times10^{-3}, 1)$ |

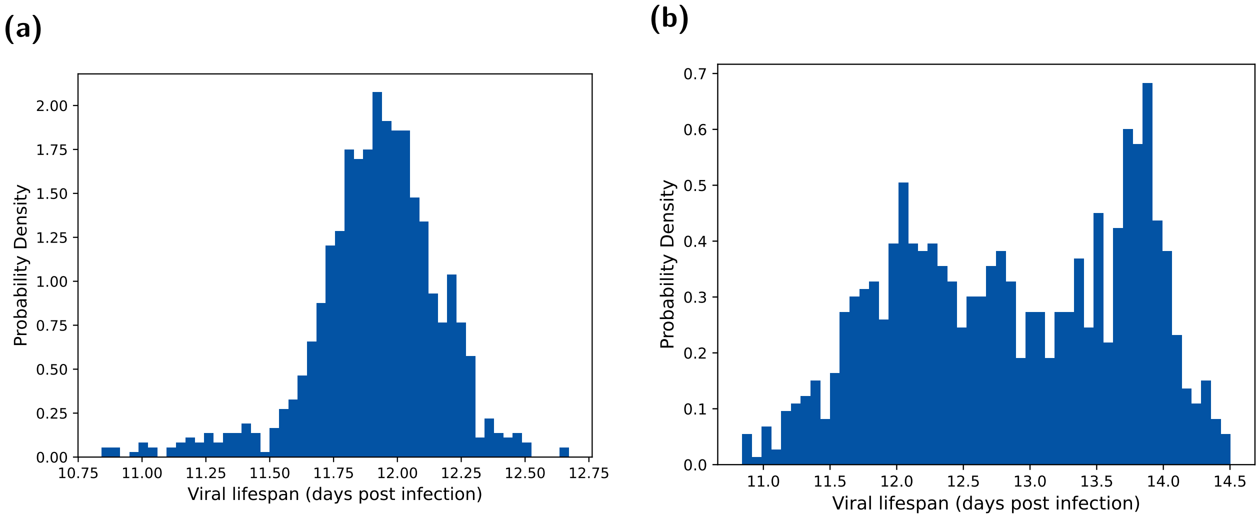


**Fig. 3. Viral lifespan ($T$) distributions under each case fatality rate assumption.** The assumed case fatality rates were **(a)** 73% and **(b)** 20%, respectively. We obtained the viral lifespan distributions by determining when either the viral load dropped below $10^4$ or the integral under the log curve reached a value $M$. We performed the fitting method outlined in Section 2.1.6.

#### 2.2 Mutation modelling and viral dynamics

From outputs that could be generated from our two-patch within-host model, we next needed an additional modelling component that would enable us to calculate the proportion of virions with zero, one, two, three, four and five mutations, and the probability that any given virion within the body had this number of mutations. In this section we outline our adapted stochastic branching process mutation model used for this purpose. This model contains biologically informed values for key model parameters, informed by the incorporation of results from the within-host infection model, thus providing a prominent modelling advance.

We adapted the stochastic branching process mutation model for viral mutation introduced in Russell *et al.* [5], in which viral replication occurs at fixed time intervals of length $\Delta$ with a mutation rate $\mu$ and replication rate $r$. The total number of viruses with $j$ mutations at each time step $t_k = k\Delta$ (with $k \in \mathbb{N}$ and $t_k < T$), $N_{t_k}^{(j)}$, is then given as a Poisson random variable:

$$N_{t_k}^{(j)} \sim \mathrm{Poi}(r \sum_{i=0}^{5} N_{t_{k-1}}^{(i)} \mu_{ij})$$

where

$$\mu_{ij} = \mathbb{P}(\text{Mutates from } i \text{ to } j \text{ mutations}) = \begin{cases} \mu^{j-i} \text{ for } i < j \\ 1 - \mathbb{1}_{\{\mathrm{i} < 5\}} \sum_{j=i+1}^{5} \mu^{j-i} \text{ for } i = j \\ 0 \text{ otherwise} \end{cases}$$

Note that the rate of $N_{t_k}^{(j)}$ is a summation due to the additive property of the Poisson random variable.

We then modified the above process to allow model parameters to be informed from the fitted within-host infection model. We allowed $r$ to be a function of time, $r(t_k)$, rather than a fixed value. Our branching process was thereby defined by:

$$N_{t_k}^{(j)} \sim \mathrm{Poi}(r(t_k) \sum_{i=0}^{5} N_{t_{k-1}}^{(i)} \mu_{ij})$$

The function $r(t_k)$ represents the viral replication rate as derived from our two-patch within-host model. To calculate this, we need the growth rate of new virions and the death rate of existing virions.

We first express a partition $P$ of $[t_k, t_{k+1}]$ such that $t_k = \tau_0 < \tau_1 < \cdots < \tau_m = t_{k+1}$ with $\tau_{i+1} - \tau_i = \delta$ where $\delta$ is the rate at which the ODE system is updated when solved numerically.

Taking $S \in \{U, L\}$, to denote whether the value corresponds to the URT or LRT, and ~~$N, V^U, D, p, I, c,$~~ $N, V^U, p, I, c,$ $V^L, \beta, T, k, X$ and $V$ to be as described in Section 2.1.3, we now define the growth rate of new virions at time $\tau_l$:

$$R_{\tau_l}^{(S)} = 1 + \frac{N_{\tau_l}^{(S)} - N_{\tau_{l-1}}^{(S)}}{V_{\tau_{l-1}}},$$

where $N^{(S)}$ solves the following differential equation for the rate of virion production:

$$\frac{dN^{(S)}}{dt} = p^{(S)} I^{(S)},$$

and we define the death rate of existing virions at time $\tau_l$:

$$K_{\tau_l}^{(S)} = 1 - \frac{D_{\tau_l}^{(S)} - D_{\tau_{l-1}}^{(S)}}{V_{\tau_{l-1}}}, K_{\tau_l}^{(S)} = 1 - \frac{Q_{\tau_l}^{(S)} - Q_{\tau_{l-1}}^{(S)}}{V_{\tau_{l-1}}},$$

where $D^{(S)}$ $Q^{(S)}$ solves the following differential equation for the rate of virion removal

$$\frac{dD^{(S)}}{dt} = cV^{(S)} + \beta^{(S)}T^{(S)}V^{(S)} + \mathbb{1}_{\{S=L\}}kXV^{(S)} \frac{dQ^{(S)}}{dt} = cV^{(S)} + \beta^{(S)}T^{(S)}V^{(S)} + \mathbb{1}_{\{S=L\}}kXV^{(S)}.$$

Our viral repilication rate $r(t_k)$ is then given by the product of the weighted sum of the number of virions created and destroyed at each time step $\delta$ in each tract:

$$r(t_k) = \prod_{l=1}^{m} \frac{\sum_{S\in\{U,L\}} R_{\tau_l}^{(S)} K_{\tau_l}^{(S)} V_{\tau_{l-1}}^{(S)}}{V_{\tau_l}}$$

$$\mathbb{P}(\text{An individual at time } t \text{ has at least one virion which has undergone } i \text{ mutations})$$
$$= 1 - \mathbb{P}(A)$$
$$= 1 - \mathbb{P}(\textstyle\bigcap_{k=1}^{V_t} B_k)$$
$$= 1 - \textstyle\prod_{k=1}^{V_t} \mathbb{P}(B_k \mid \bigcap_{j<k} B_j)$$
$$\leq 1 - \textstyle\prod_{k=1}^{V_t} \mathbb{P}(B_k)$$
$$= 1 - \textstyle\prod_{k=1}^{V_t} (1 - \frac{V_t^{(i)}}{V_t})$$
$$= 1 - (1 - \frac{V_t^{(i)}}{V_t})^{V_t} = \hat{p}_t^{(i)}, i = 0, 1, 2, 3, 4, 5$$

To justify the inequality, we first note that if an arbitrary virion at the current timestep has $i$ mutations, the probability that any other virion has that number of mutations would increase. This is because there is a chance that virions with the same number of mutations could have the same parent. The joint probability events $(B_k \mid \bigcap_{j<k} B_j)$ takes into account this positive correlation (but the event $B_k$ by itself does not), i.e. $\mathbb{P}(B_k \mid \bigcap_{j<k} B_j) \geq \mathbb{P}(B_k)$

Additionally, since each virion is equally likely to mutate, we used the proportion of virions with $i$ mutations to get that $\mathbb{P}(B_k) = 1 - \frac{V_t^{(i)}}{V_t} \quad \forall k$.

### 3 Results

#### 3.1 Fitting the two-tract within-host respiratory infection model to H5N1 influenza viral titre data

Having developed our within-host respiratory infection model, with infection dynamics in the LRT and URT modelled explicitly, it was important to ascertain whether it could reproduce the observed H5N1 influenza viral titres (Fig. 1) whilst maintaining biologically reasonable parameters. Resultant parameter posteriors would then be used as inputs to the branching process model.

We ran the ABC-SMC-MNN routine and obtained 1000 samples of the posterior distribution for seven fitted parameters: $\beta_U$, $\beta_L$, $p_U$, $p_L$, $\gamma$, $D$ and $a$ (Fig. 4). We note that even for the posteriors that had similarities to a log-uniform distribution ($\beta_L, p_L, \gamma, D, a$), the range and probability mass of these distributions shifted compared to the prior. This is reinforced by a least-squares fit producing a similar profile to the median of the posterior-predictive distribution (Fig. 5(a)). The least-squares fit parameters can be found in Section S1.

Comparing the inferred posterior distributions for the URT and LRT spreading rate parameters, the 95% credible interval for the spreading rate in the URT ($\beta_U \in [2.24\times10^{-7}, 3.47\times10^{-7}]$) was generally at a lower range than in the LRT ($\beta_L \in [1.09\times10^{-7}, 1.18\times10^{-6}]$). This difference possibly corresponds to the preferential binding of H5N1 influenza to the epithelial cells in the LRT than in the URT. The production rate in the URT ($p_U \in [0.348, 0.608]$) was higher than in the LRT ($p_L \in [0.00488, 0.044]$), likely due to the higher target-cell count (and thus maximum production rate) in the LRT. There was a clear negative correlation between $\beta_U$ and $p_U$ (relating to the previous discussion), which is to be expected as an increase in the spreading rate would lead to target-cells being infected sooner and hence a larger infection time available to produce virions (meaning that a lower $p_U$ is required) and vice-versa. The 95% credible interval for $\gamma$ was at a low range of $[2.72\times10^{-6}, 2.97\times10^{-4}]$, indicating that the parameter was needed to delay the peak time, but only at smaller values. The 95% credible intervals for the diffusion ($D \in [0.00141, 0.0680]$) and advection ($a \in [0.0414, 0.731]$) coefficients are quite wide, possibly indicating that the intra-patch processes contribute more than the inter-patch processes to the viral dynamics.

Through simulation of our model using the 1000 parameter sets representing samples from the target posterior distribution, we next checked the correspondence between the posterior predictive distribution for $V_U$ and the empirical viral titre data (Fig. 5(a)). As the data is wide ranging - the smallest viral titre measured had a value of around $10^4 TCID_{50}/ml$ with the highest being at over $10^8$ - the likelihood surface has many steep local minima giving a tight posterior interval. However, it can also be seen that the least squares solution (Section S1) lies close to the median produced by the ABC method, giving confidence about the validity of the solution. The predictive interval lay within the middle range of the dataset. The qualitative shape (including peak height and time) of the median was very similar to other models [6, 13, 18, 36]. For the least-squares optimisation we chose five different starting parameter sets and selected the resultant local mode with the lower error. Although not guaranteed, we are confident that this is likely the global optima as multiple starting points converged to this value. This outcome supports the parameter posterior distributions acquired by the ABC approach successfully incorporating the posterior.

Lastly, inspection of the peak time distribution of viral titre realisations from the posterior predictive distribution showed almost all of the density of peak viral titre occurring between 4.7 and 5 days post infection (Fig. 5(c)). This observation provided further assurance in the concordance between the fitted model and the empirical data.

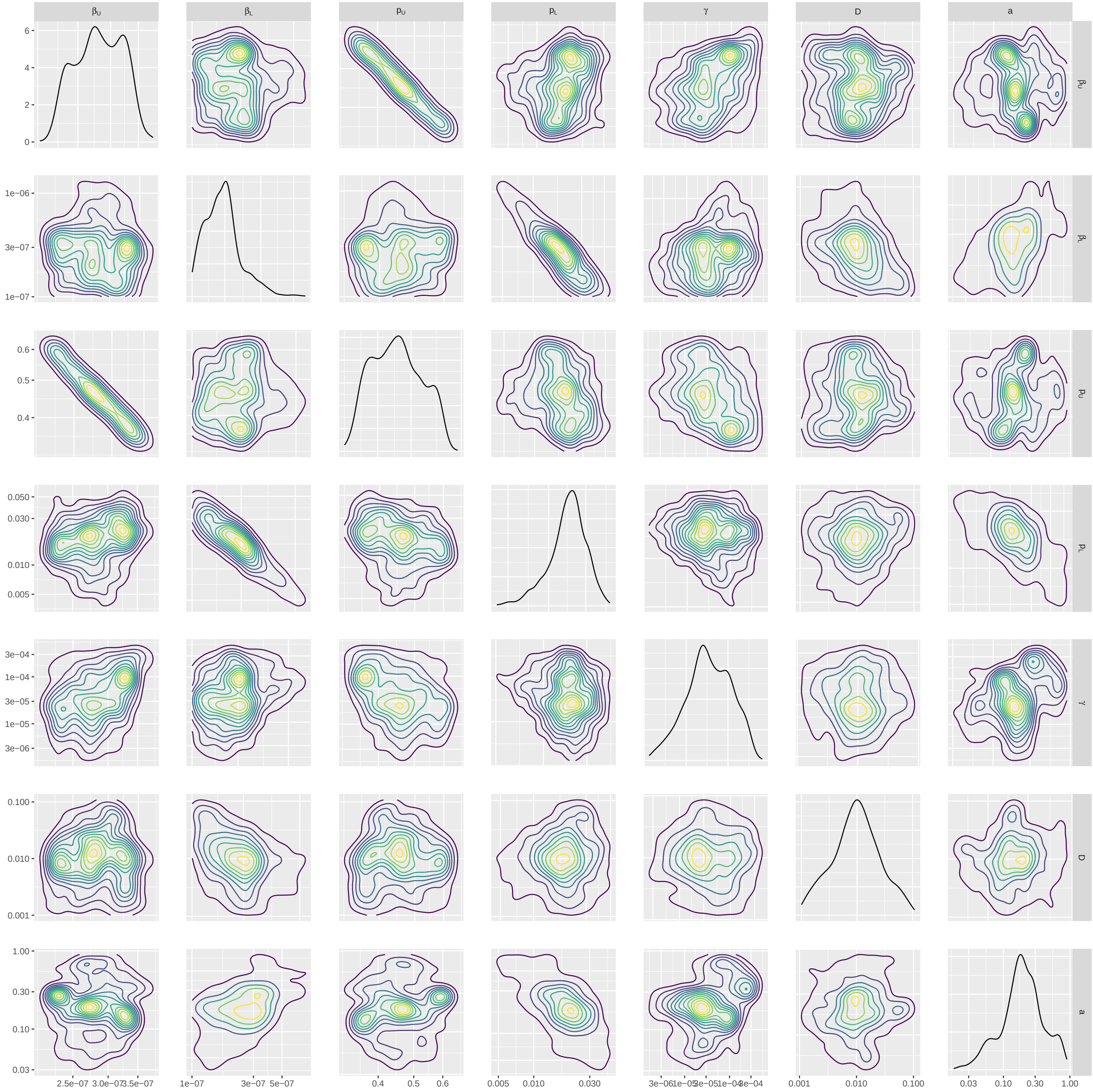


**Fig. 4. Parameter posterior distributions.** We obtained 1000 samples of the target posterior distribution using the ABC-SMC-MNN method outlined in Section 2.1.5. Diagonal panels show the marginal distributions for: rate of infection in the URT ($\beta_U$) and the LRT ($\beta_L$), virus reproduction rate in the URT ($p_U$) and the LRT ($p_L$), viral uptake rate between infection and viral titre ($\gamma$), rate of diffusion of free virions ($D$) and the rate of advection ($a$), respectively. Off-diagonal panels show bi-parameter distributions, where the contour shading intensity corresponds to the probability density value (lighter for higher probability density). Parameters ($\beta_L, p_L$) in the LRT tended to be higher than the URT ($\beta_U, p_U$), agreeing with the biological preference for H5N1 influenza to infect the LRT.

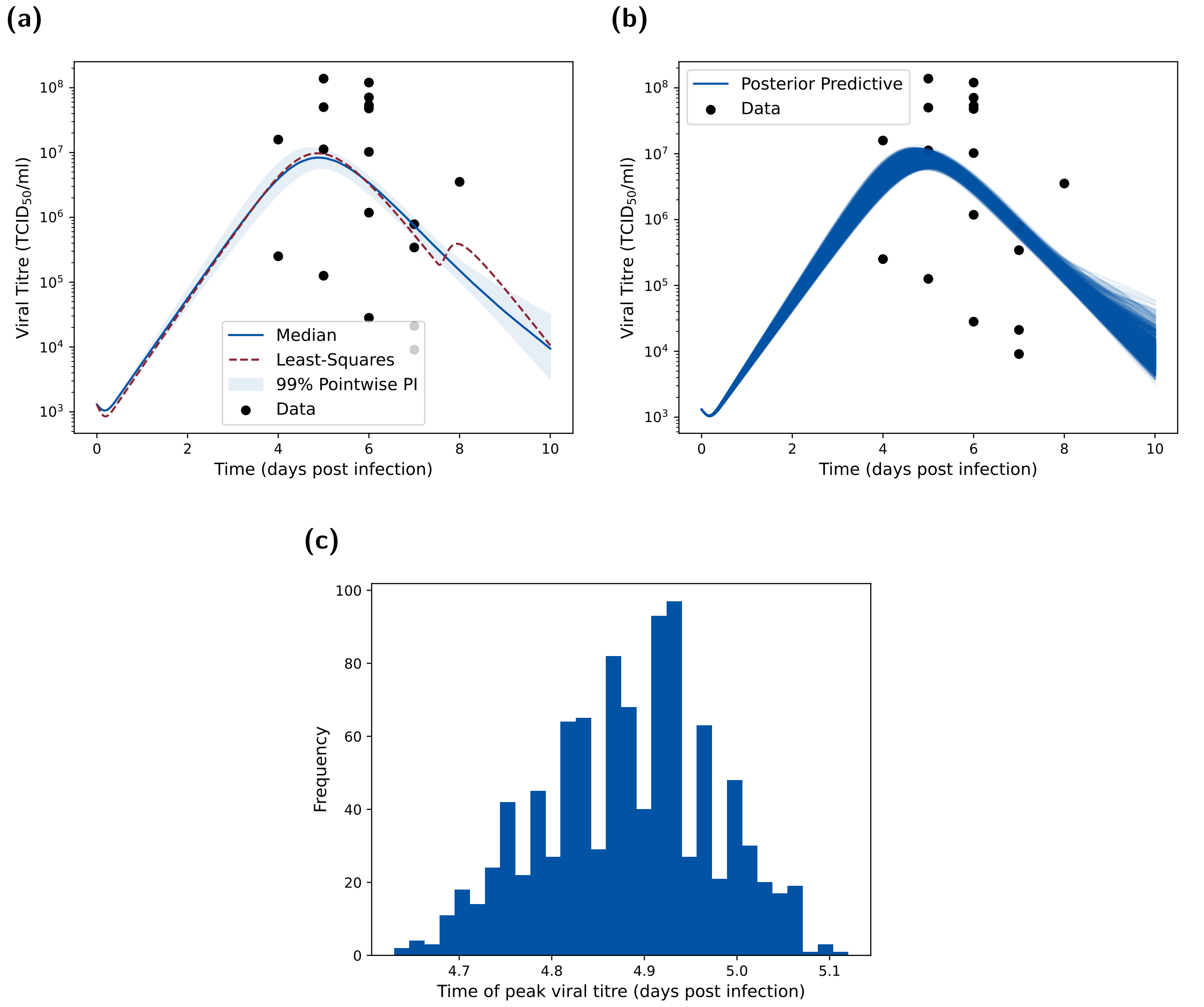


**Fig. 5. Posterior predictive distributions for URT viral titre metrics. (a)** Posterior predictive distribution for $V_U$ compared to the empirical data. We produced the posterior predictive distribution using the 1000 parameter samples from our inferred parameter posterior distribution in Fig. 4. We display the median (blue solid line), 95% pointwise prediction interval (shaded region) and the least-squares fit (dotted red line). Both the optimisation fit and ABC posterior show reasonable concordance to the main data trends. **(b)** As for **(a)**, but showing all posterior predictive trajectories as opposed to the distribution summary. **(c)** Posterior viral titre peak-time distribution showing that the majority of infections peak just before day five. This is consistent with the data, which shows a peak around day five to day six (post infection).

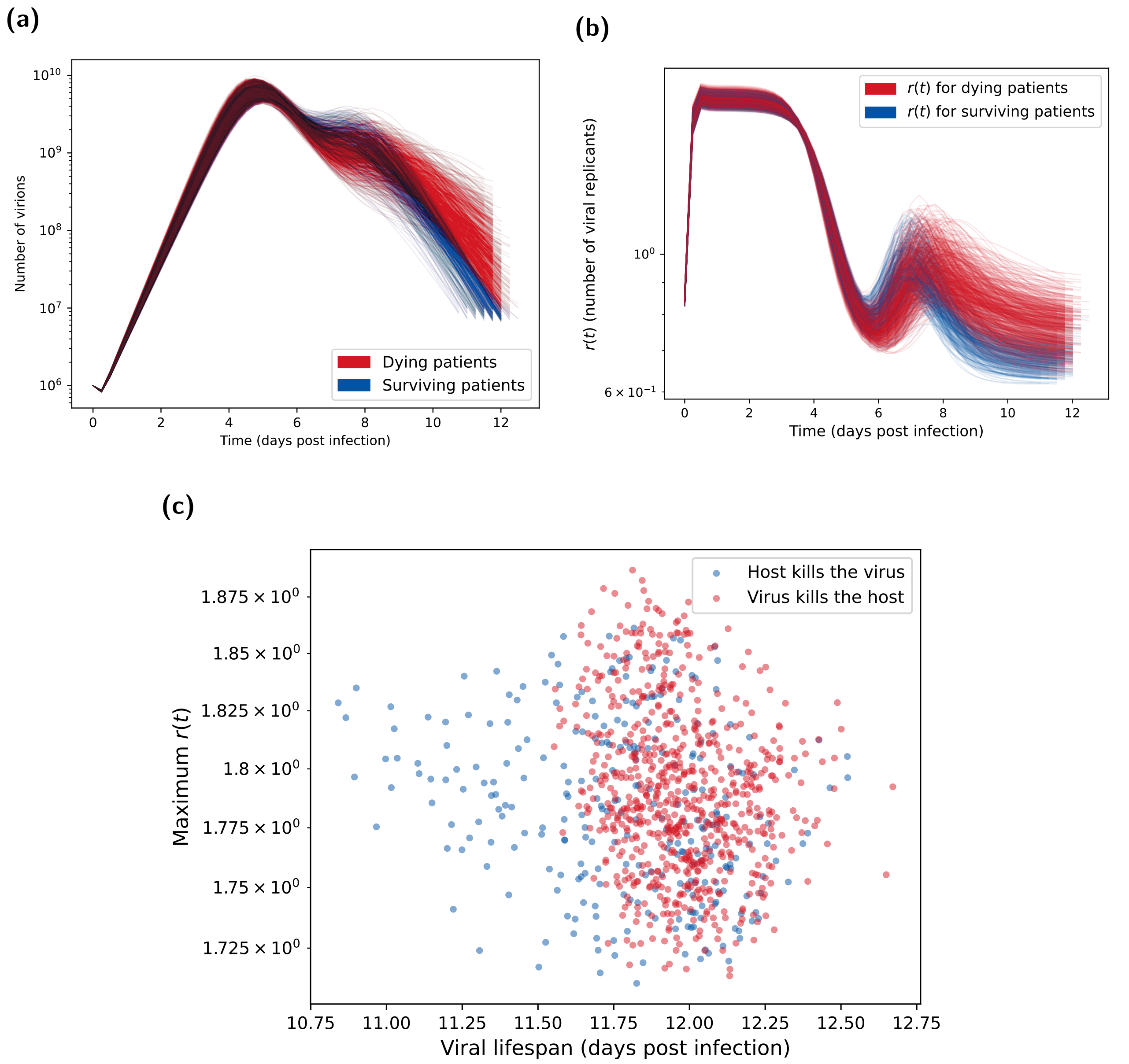


**Fig. 6. Posterior predictions for number of virions and number of viral replicants ($r(t)$).** Both plots show the 1000 posterior trajectories, with the blue lines representing H5N1 influenza patients who survive the infection (cleared the virus) and the red lines representing patients who died due to the infection (where the distinction is made using the method in Section 2.1.6). **(a)** Virion count distribution found using the parameter posterior in Fig. 4. After day 6, the viral count trajectories for deceased patients are higher and more sustained than surviving patients. These were calculated from the one million BPM realisations each with an initial viral load of $10^6$ and a mutation chance of $10^{-5}$ per replication. **(b)** Distribution of number of viral replicants ($r(t)$) calculated from the posterior predictive distribution shown in Fig. 5(a). Surviving patients tended to have higher $r(t)$ during the second peak around day 7, which then declined below one (indicating a decreasing virion count) more rapidly than for dying patients. **(c)** Maximum $\boldsymbol{r}$ value vs viral lifespan. Maximum $r$ values taken from Fig. 6(b) and corresponding viral lifespans are shown in Fig. 3. Blue circles represent H5N1 influenza patients who survive the infection (cleared the virus). Red circles represent patients who died due to the infection (where the distinction is made using the method in Section 2.1.6). Surviving individuals tended to have shorter viral lifespans. Otherwise, there was no evident dependencies between maximum $r$ value and the viral lifespan.

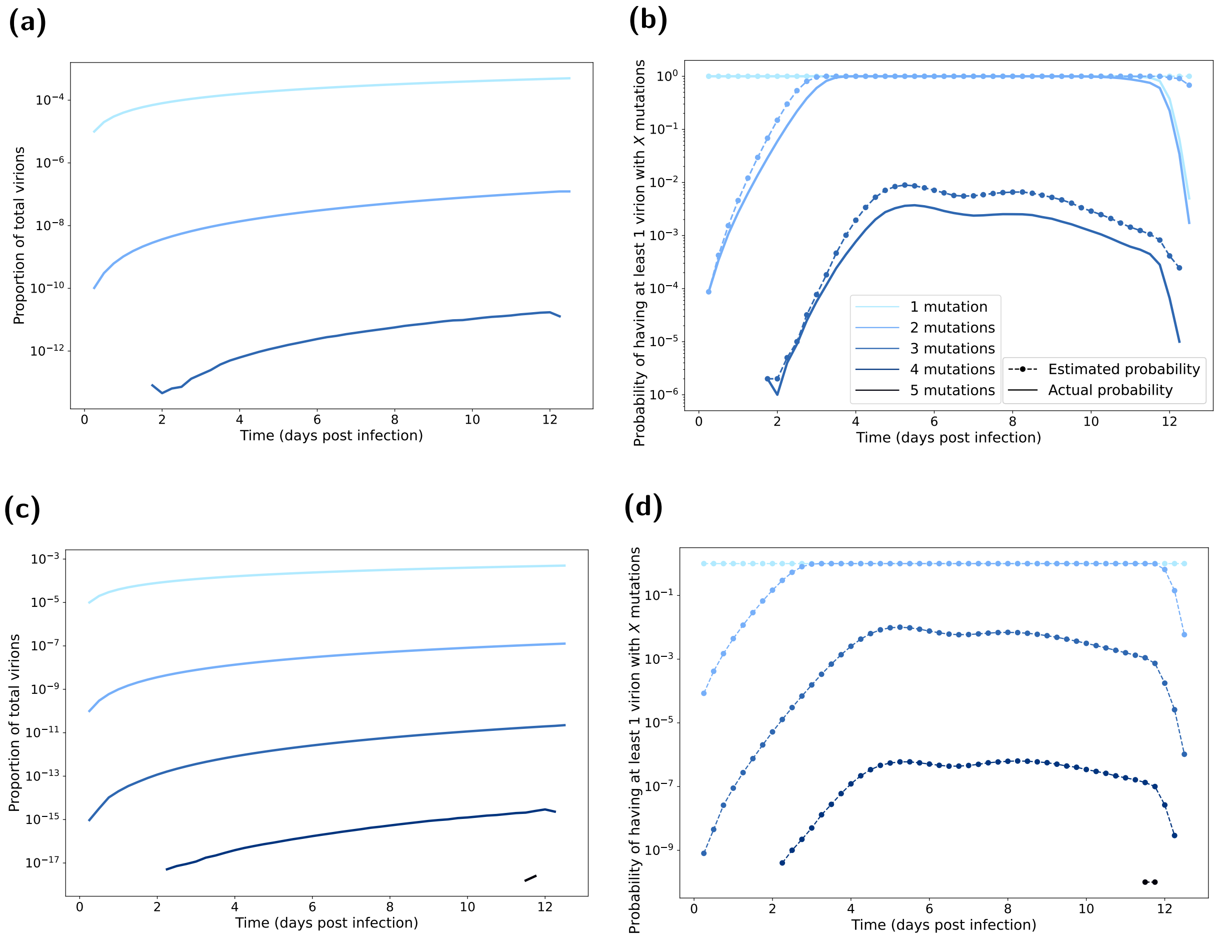


**Fig. 7. Mutated virion statistics with respect to time elapsed post infection, computed from BPM realisations.** Line shading corresponds to the number of mutations (one mutation the lightest shading through to five mutations being the darkest shading). In all BPM simulations we fixed the probability of mutation at $10^{-5}$ per replication. In panels **(a&b)**, each realisation had an initial viral load of $10^6$. We ran 1000 realisations of each of the 1000 posterior parameter sets (Fig. 4). In panels **(c&d)**, each realisation had an initial viral load of $10^6 \times 10^6$. We ran one realisation of each of the 1000 posterior parameter sets (Fig. 4). **(a,c)** Proportion of total virions with the specified amount of mutations. There were a very small proportion of virions that have the required number of mutations to achieve droplet transmission (three or more mutations). **(b,d)** Probability of having a mutation strain. We present the estimated probabilities as the dashed lines with circle markers. We present the actual probabilities as solid lines. Probability estimate derivation follows that given in Section 2.2. The estimated probabilities are generally a clear upper bound on the true probabilities. Although not seen, we expect the only exceptions to be for higher number of mutations, where estimated probabilities for when these strains first occur could be initially below the actual probability; this reflected the dependence on the population of other mutants being more pronounced at lower numbers of virions (where the presence of a four mutation strain, for example, is almost purely from mutations from zero, one, two, three strain virions). Depending on the number of mutations in the initial infecting virions, there was a low probability of achieving the required number of mutations near the beginning on the infection lifespan (which would allow more replication of the mutant strains).

Archived code available at:
`https://doi.org/10.5281/zenodo.13385415`.

#### Competing interests

All authors declare that they have no competing interests.

#### Funding statement

DH, LJ, RS and MJK were supported by the Engineering and Physical Sciences Research Council through the MathSys CDT [grant number EP/S022244/1]. MJK was also supported by the Medical Research Council through the JUNIPER partnership award [grant number MR/X018598/1]. EMH is also a member of the JUNIPER partnership (MRC grant no MR/X018598/1) and would like to acknowledge their help and support. EMH is affiliated to the National Institute for Health and Care Research Health Protection Research Unit (NIHR HPRU) in Gastrointestinal Infections at University of Liverpool (PB-PG-NIHR-200910), in partnership with the UK Health Security Agency (UKHSA), in collaboration with the University of Warwick (EMH is based at The University of Liverpool). The views expressed are those of the author(s) and not necessarily those of the NIHR, the Department of

Health and Social Care or the UK Health Security Agency. The research was funded by The Pandemic Institute, formed of seven founding partners: The University of Liverpool, Liverpool School of Tropical Medicine, Liverpool John Moores University, Liverpool City Council, Liverpool City Region Combined Authority, Liverpool University Hospital Foundation Trust, and Knowledge Quarter Liverpool (EMH is based at The University of Liverpool). The views expressed are those of the author(s) and not necessarily those of The Pandemic Institute. The funders had no role in study design, data collection and analysis, decision to publish, or preparation of the manuscript. For the purpose of open access, the authors have applied a Creative Commons Attribution (CC BY) licence to any Author Accepted Manuscript version arising from this submission.

#### ~~Data availability~~

~~All data utilised in this study are publicly available, with relevant references and data repositories provided.~~

#### ~~Code availability~~

~~The code repository for the study is available at:~~

### Supporting Information
### Introducing a framework for within-host dynamics and mutations modelling of H5N1 influenza infection in humans

Daniel Higgins[1,2‡*], Joshua Looker[1,2‡*], Robert Sunnucks[1,2‡*], Jonathan Carruthers[3], Thomas Finnie[3], Matt J. Keeling[2,4], Edward M. Hill[5,6*]

~~Archived code available at:~~ **1** EPSRC & MRC Centre for Doctoral Training in Mathematics for Real-World Systems, University of Warwick, Coventry, United Kingdom.

#### ~~Competing interests~~

~~All authors declare that they have no competing interests.~~ **2** The Zeeman Institute for Systems Biology & Infectious Disease Epidemiology Research, University of Warwick, Coventry, United Kingdom.
**3** Data, Analytics and Surveillance, UK Health Security Agency, London, United Kingdom.
**4** Mathematics Institute and School of Life Sciences, University of Warwick, Coventry, United Kingdom.
**5** Civic Health Innovation Labs and Institute of Population Health, University of Liverpool, Liverpool, United Kingdom.
**6** NIHR Health Protection Research Unit in Gastrointestinal Infections, University of Liverpool, Liverpool, United Kingdom.

‡These authors contributed equally to this work.

#### Table of Contents

**S1 Additional tables** 2

**S2 Additional figures** 2

**S3 Robustness of model outcomes when applying an incubation period to viral load data** 2

**S4 Robustness of model outcomes to a lower case fatality rate** 8

### S1 Additional tables

**Table S1. Least-squares fit parameters:** and their descriptions. Parameters fitted to the data in the log-space using the 'scipy.optimize' library in Python. We present values under two different assumptions for mapping days post-infection in the model to the de Jong *et al.* [1] viral load data that is dependent on 'days since onset of illness'. We amend the de Jong *et al.* [1] viral load data to be in terms of days post-infection rather than 'days since onset of illness' by applying either no latent period or a 5 day latent period (LP).

| **Parameter** | | **Value** – no LP | **Value** – 5 day LP |
|---|---|---|---|
| $\beta_U$ | Rate of infection, URT | $3.027 \times 10^{-7}$ | $1.870 \times 10^{-7}$ |
| $\beta_L$ | Rate of infection, LRT | $9.59 \times 10^{-4}$ | $2.211 \times 10^{-6}$ |
| $p_U$ | Virus production rate, URT | 0.588 | 0.524 |
| $p_L$ | Virus production rate, LRT | $7.71 \times 10^{-2}$ | 0.140 |
| $\gamma$ | Infectious virions and $TCID_{50}$ / PFU conversion | $1.16 \times 10^{-2}$ | $2.520 \times 10^{-2}$ |
| $D$ | Rate of diffusion of free virions | 0.215 | $7.858 \times 10^{-3}$ |
| $a$ | Rate of advection | $1.95 \times 10^{-2}$ | 0.150 |

### S2 Additional figures

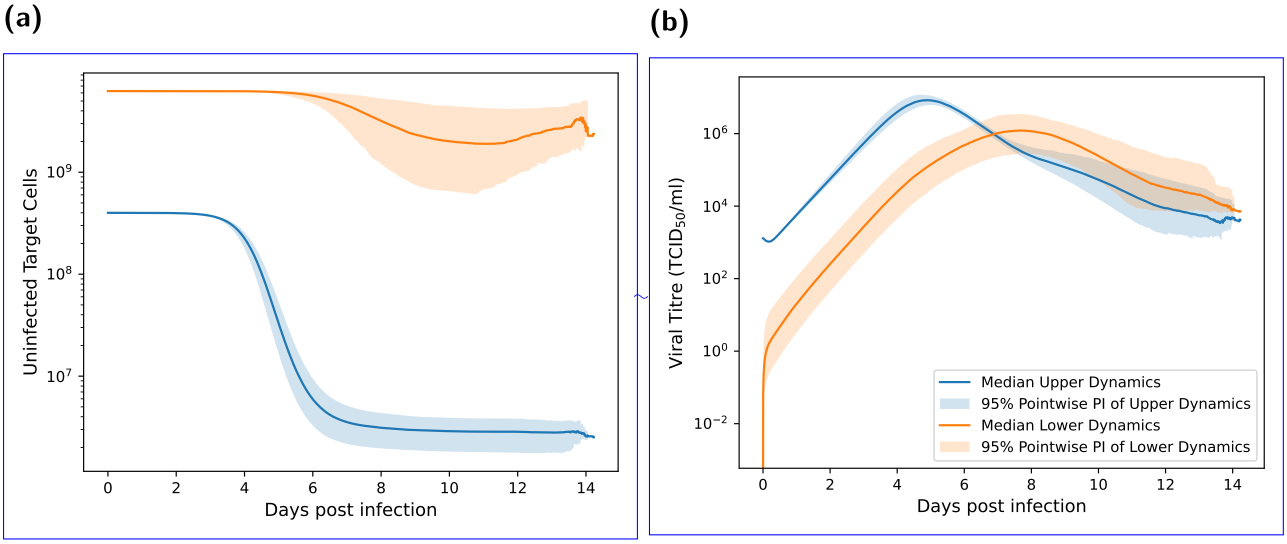


**Fig. S1. Compartmental trajectories for uninfected target cells and free virions.** Both plots show the median and 95% pointwise predictive interval of the 1000 posterior trajectories, with the blue lines representing dynamics of the URT and the orange lines representing dynamics of the LRT. **(a)** The target cell count trajectories. Most of the target cells in the URT are killed by virions, with the number of uninfected target cells dropping by two orders of magnitude. In the LRT, the number of target cells still decreases, but not to the extent as seen in the URT. **(b)** The free virion's viral titre trajectories. Viral trajectories peak higher and earlier in the URT, at around $10^7 TCID_{50}$/ml at approximately four days post infection. Peaks in viral trajectories for the LRT are lower and occurred later, at roughly $10^6 TCID_{50}$/ml after 7-8 days post infection.

### S3 Robustness of model outcomes when applying an incubation period to viral load data

Here we present results assuming an incubation period of five days when calibrating model outputs in terms of days post-infection to the de Jong *et al.* [1] viral load data that was dependent on 'days since onset of illness'. An incubation period of five days equals the median incubation period reported by a retrospective cohort study of 18 influenza virus (H5N1) cases in China [2]. Compared to our main analysis findings, under this alternative incubation period assumption we observed a similar posterior distribution for $\beta_U, p_U$ and $\gamma$ (Fig. S2). Although the $\beta_L$ has most of its distribution mass much higher than $\beta_U$, and $p_L$ lower than $p_U$, we note that the differences in order of magnitude between them have decreased. The rate of diffusion is now much smaller and the rate of advection much higher. These changes are likely due to the five day latent period requiring delayed and possibly smaller take-off in the lower respiratory tract than the results assuming a zero day incubation period.

For the posterior distribution of replication rates ($r(t)$) trajectories, in the majority of realisations $r(t)$ between days zero and six post infection was essentially constant, corresponding to the exponential growth of $v$ (Fig. S3). In contrast to the main analysis, there was not a second increase in replication rates. Instead, all replication rate trajectories had a consistent decline until about day 12 post infection, beyond which they plateaued (Fig. S3(b)).

Studying the relationship between viral lifespan and peak replication rate ($r(t)$), there was once a negative correlation between the viral lifespan and peak replication rate (Fig. S4).

Our general conclusions from the branching process model analysis also remained unchanged (Fig. S5 ).

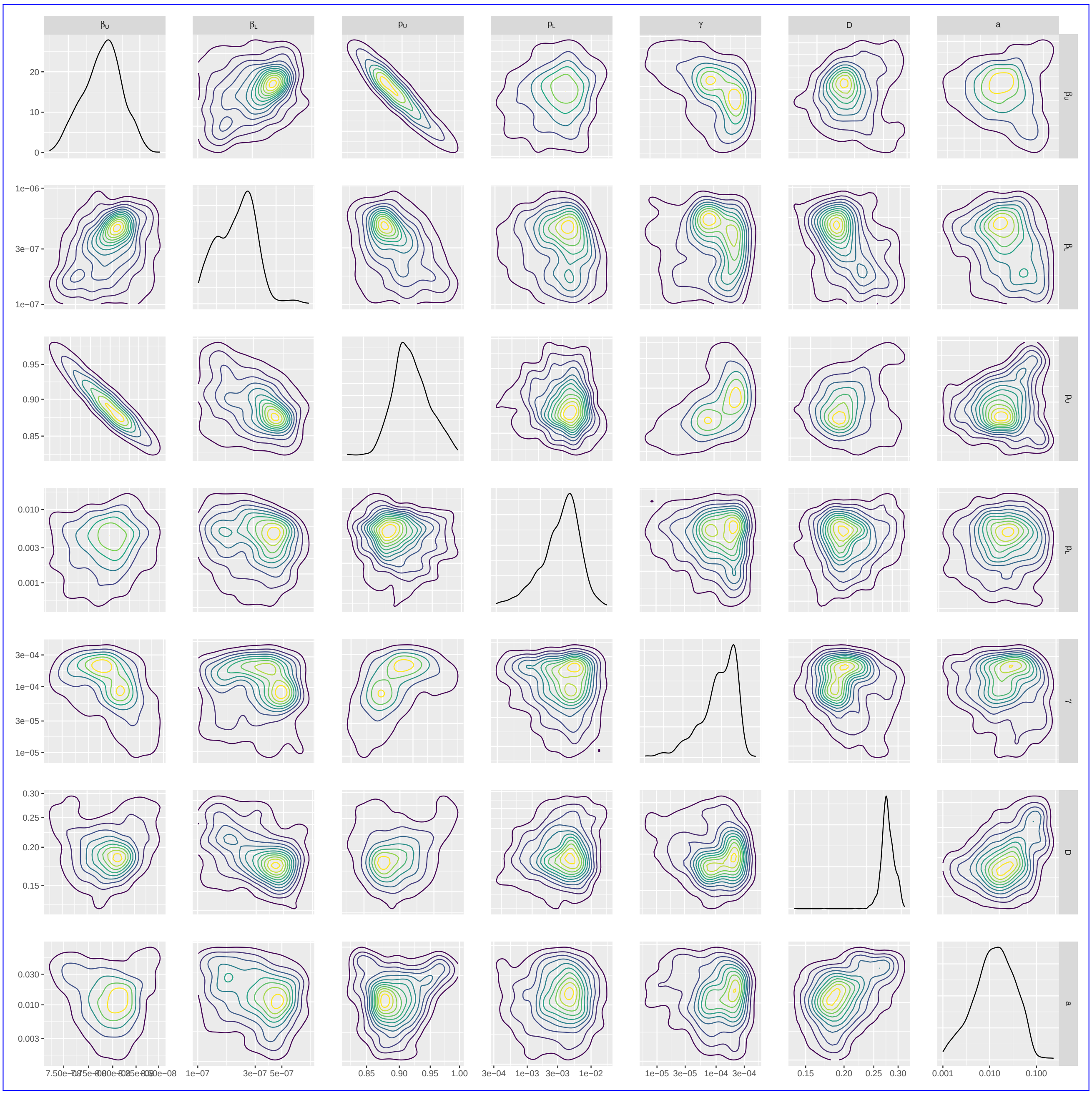


**Fig. S2.** **Parameter posterior distributions.** We obtained 1000 samples of the target posterior distribution using the ABC-SMC-MNN method outlined in Section 2.1.5. Diagonal panels show the marginal distributions for: rate of infection in the URT ($\beta_U$) and the LRT ($\beta_L$), virus reproduction rate in the URT ($p_U$) and the LRT ($p_L$), viral uptake rate between infection and viral titre ($\gamma$), rate of diffusion of free virions ($D$) and the rate of advection ($a$), respectively. Off-diagonal panels show bi-parameter distributions, where the contour shading intensity corresponds to the probability density value (lighter for higher probability density). Parameters ($\beta_L, p_L$) in the LRT tended to be higher than the URT ($\beta_U, p_U$), agreeing with the biological preference for H5N1 influenza to infect the LRT.

**(a)**

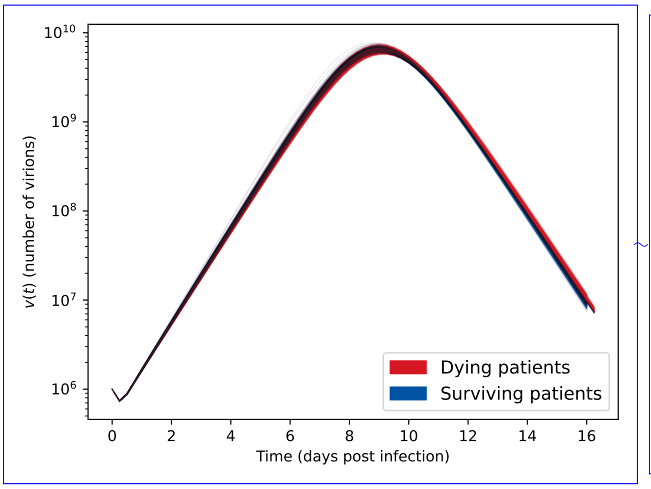


**(b)**

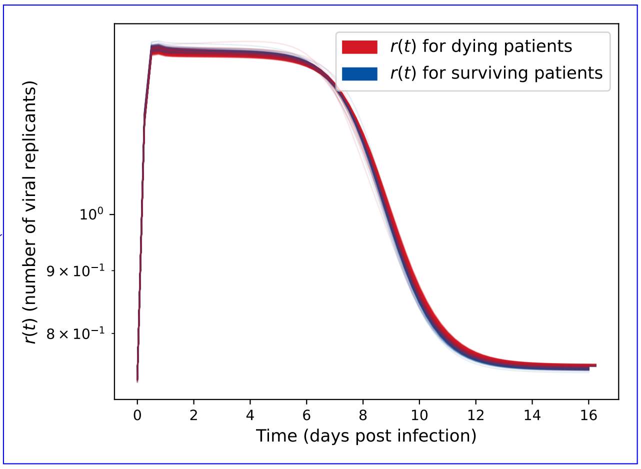


**Fig. S3.** **Posterior predictions for number of virions and number of viral replicants ($r(t)$)** assuming a five day latent period. Both plots show the 1000 posterior trajectories, with the blue lines representing H5N1 influenza patients who survive the infection (cleared the virus) and the red lines representing patients who died due to the infection (where the distinction is made using the method in Section 2.1.6). **(a)** Virion count distribution found using the parameter posterior in Fig. 4 . We see similar trends between surviving and deceased patients. We calculated these viral count trajectories from the one million BPM realisations each with an initial viral load of $10^6$ and a mutation chance of $10^{-5}$ per replication. **(b)** Distribution of $r(t)$ calculated from the posterior predictive distribution shown in Fig. 5(a). Surviving patients tended to have slightly higher $r(t)$ in the peak, which then declined below one (indicating a decreasing virion count) marginally faster than for dying patients.

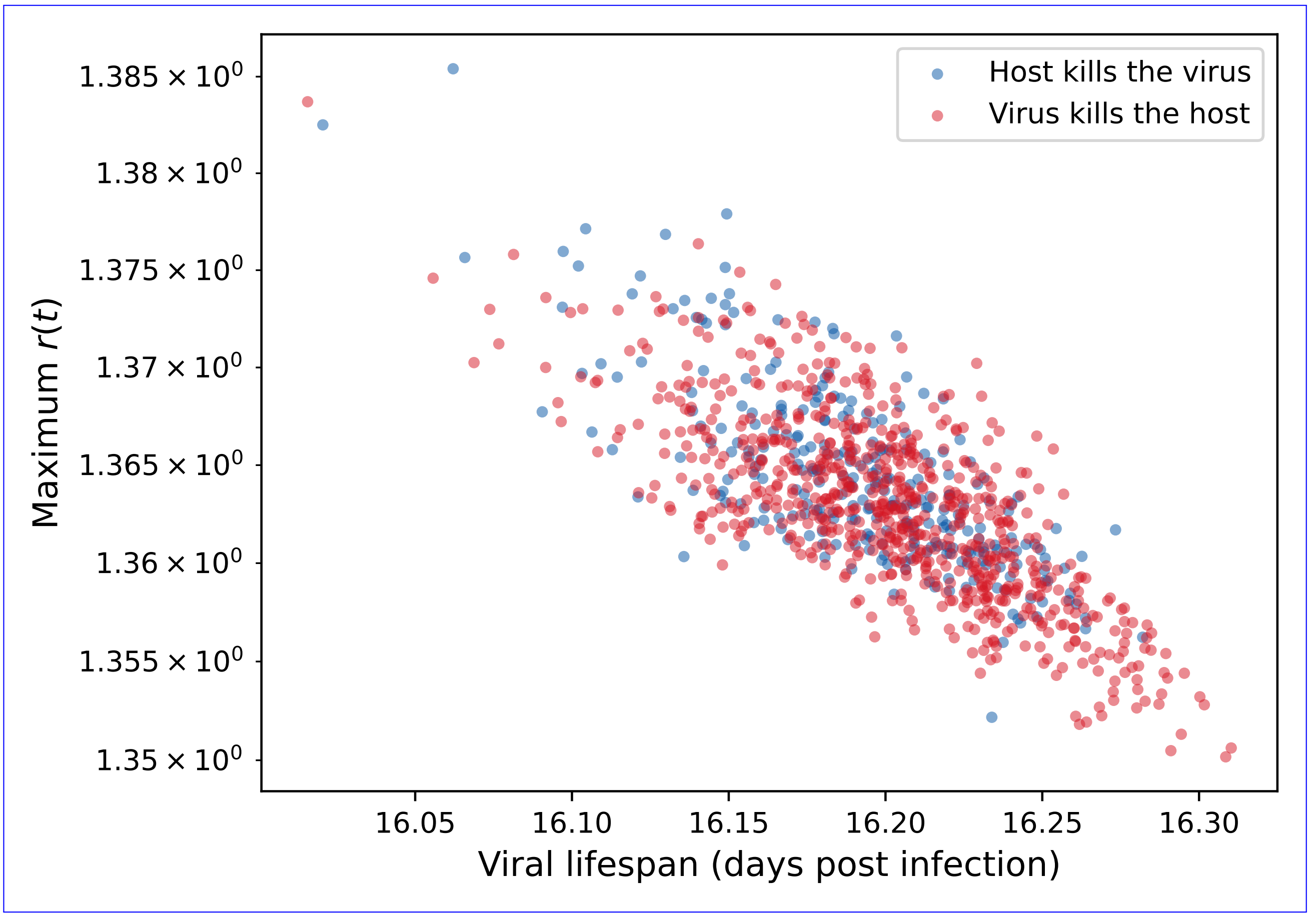


**Fig. S4.** **Maximum $r$ value vs viral lifespan assuming a five day latent period.** We observe a negative correlation between maximum $r$ value and the viral lifespan. Blue circles represent H5N1 influenza patients who survive the infection (cleared the virus). Red circles represent patients who died due to the infection (where the distinction is made using the method in Section 2.1.6). Surviving individuals tended to have lower viral lifespans.

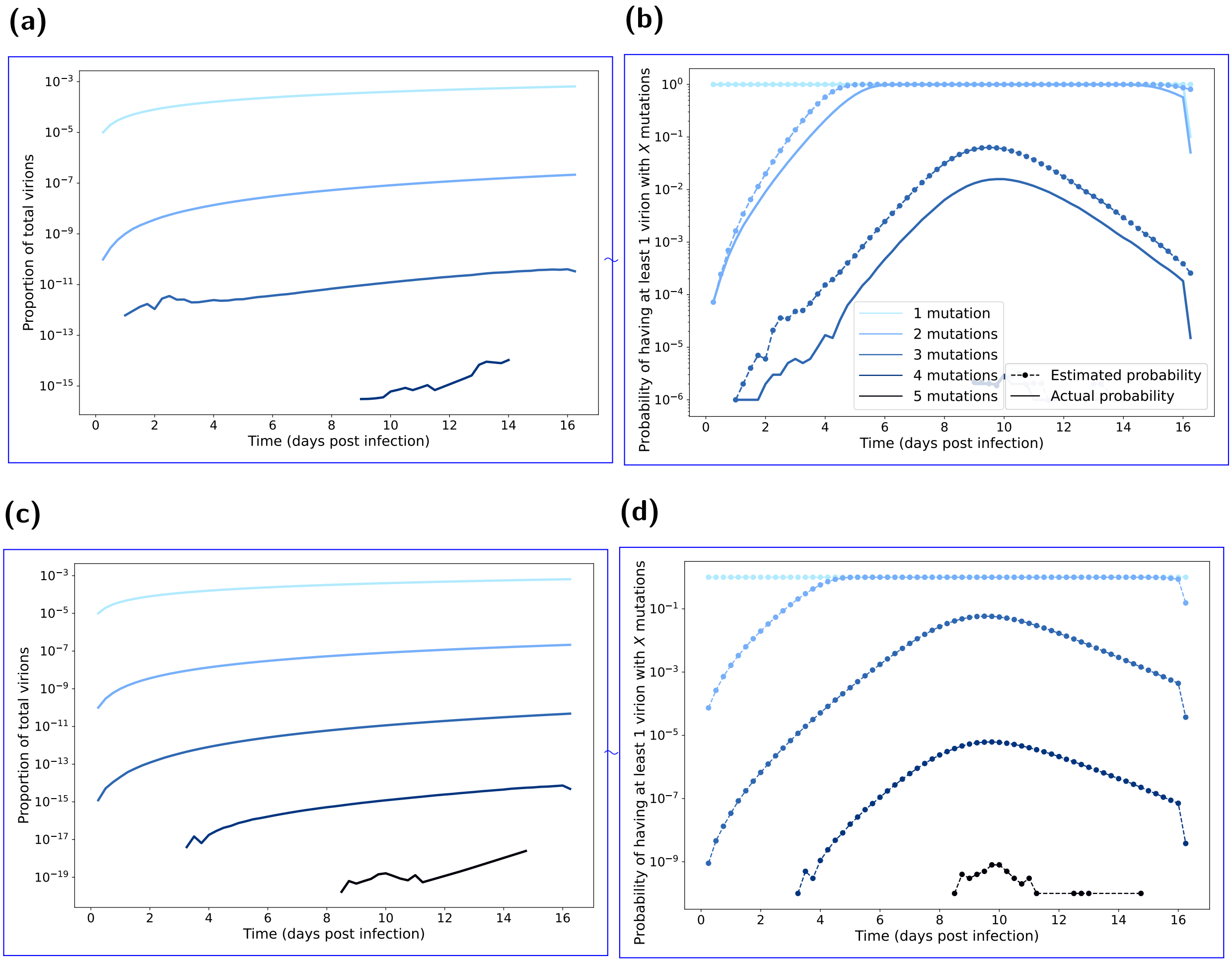


**Fig. S5.** **Mutated virion statistics with respect to time elapsed post infection, computed from BPM realisations assuming a five day latent period**. Line shading corresponds to the number of mutations (one mutation the lightest shading through to five mutations being the darkest shading). In all BPM simulations we fixed the probability of mutation at $10^{-5}$ per replication. In panels **(a&b)**, each realisation had an initial viral load of $10^6$. We ran 1000 realisations of each of the 1000 posterior parameter sets (Fig. 4). In panels **(c&d)**, each realisation had an initial viral load of $10^6 \times 10^6$. We ran one realisation of each of the 1000 posterior parameter sets (Fig. 4). **(a,c)** Proportion of total virions with the specified amount of mutations. There were a very small proportion of virions that have the required number of mutations to achieve droplet transmission (three or more mutations). **(b,d)** Probability of having a mutation strain. We present the estimated probabilities as the dashed lines with circle markers. We present the actual probabilities as solid lines. Probability estimate derivation follows that given in Section 2.2. The estimated probabilities are a clear upper bound on the true probabilities. Depending on the number of mutations in the initial infecting virions, there was a low probability of achieving the required number of mutations near the beginning on the infection lifespan (which would allow more replication of the mutant strains).

#### S4 Robustness of model outcomes to a lower case fatality rate

Here we present the results when conducting the analysis assuming a lower case fatality rate of 20%. We took the case fatality rate estimate of 20% from Dobrovolny *et al.* [3] for individuals treated with neuraminidase inhibitors.

Inspecting the viral dynamics results, with the viral trajectories being simulated before fitting to the case fatality rate, we found the viral dynamic results assuming a lower case fatality rate to be similar to the main analysis (Figs. S6 and S7).

Comparing the estimated mutation probabilities between the main analysis and when assuming the lower case fatality rate of 20%, we observed similar mutation probabilities (Fig. S8). The overall viral dynamics remained near identical for the majority of the infectious period. Consequently, the mutation probabilities were not affected until the end of the infection, by which point the peak of the mutation probabilities had already occurred.

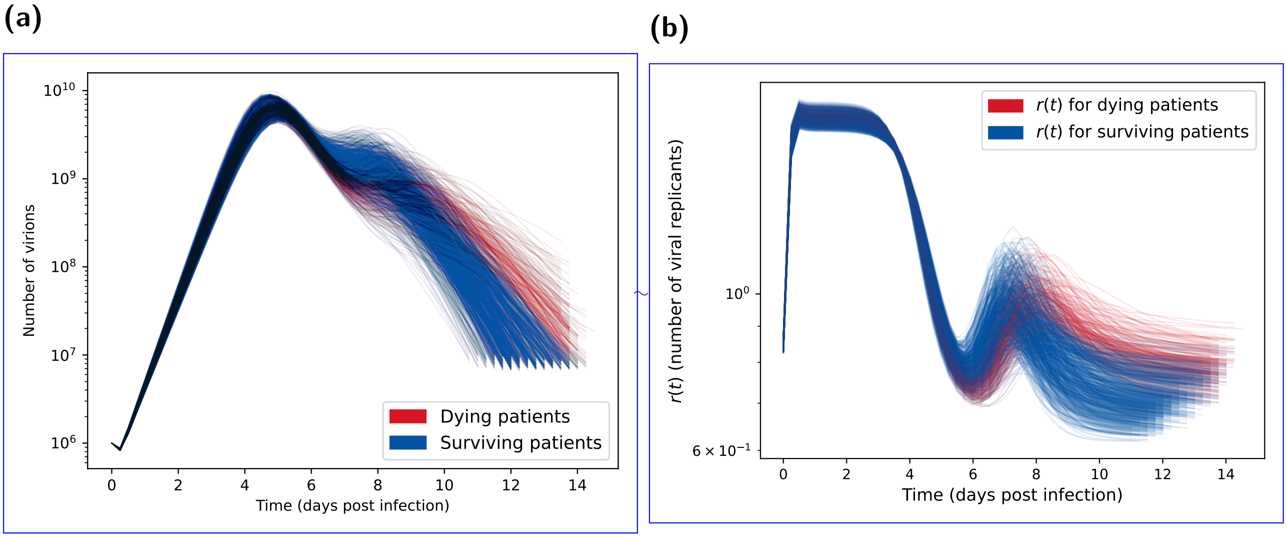


**Fig. S6. Posterior predictions for number of virions and number of viral replicants ($r(t)$) assuming a lower case mortality rate (20% instead of 73%).** Both plots show the 1000 posterior trajectories, with the blue lines representing H5N1 influenza patients who survive the infection (cleared the virus) and the red lines representing patients who died due to the infection (where the distinction is made using the method in Section 2.1.6). **(a)** Virion count distribution found using the parameter posterior in Fig. 4. The viral count trajectories for deceased patients are lower and more sustained than surviving patients. We calculated these viral count trajectories from the one million BPM realisations each with an initial viral load of $10^6$ and a mutation chance of $10^{-5}$ per replication. **(b)** Distribution of $r(t)$ calculated from the posterior predictive distribution shown in Fig. 5(a). Surviving patients tended to have higher $r(t)$ in the second peak around day 7 on infection, which then declined below one (indicating a decreasing virion count) more rapidly than for dying patients.

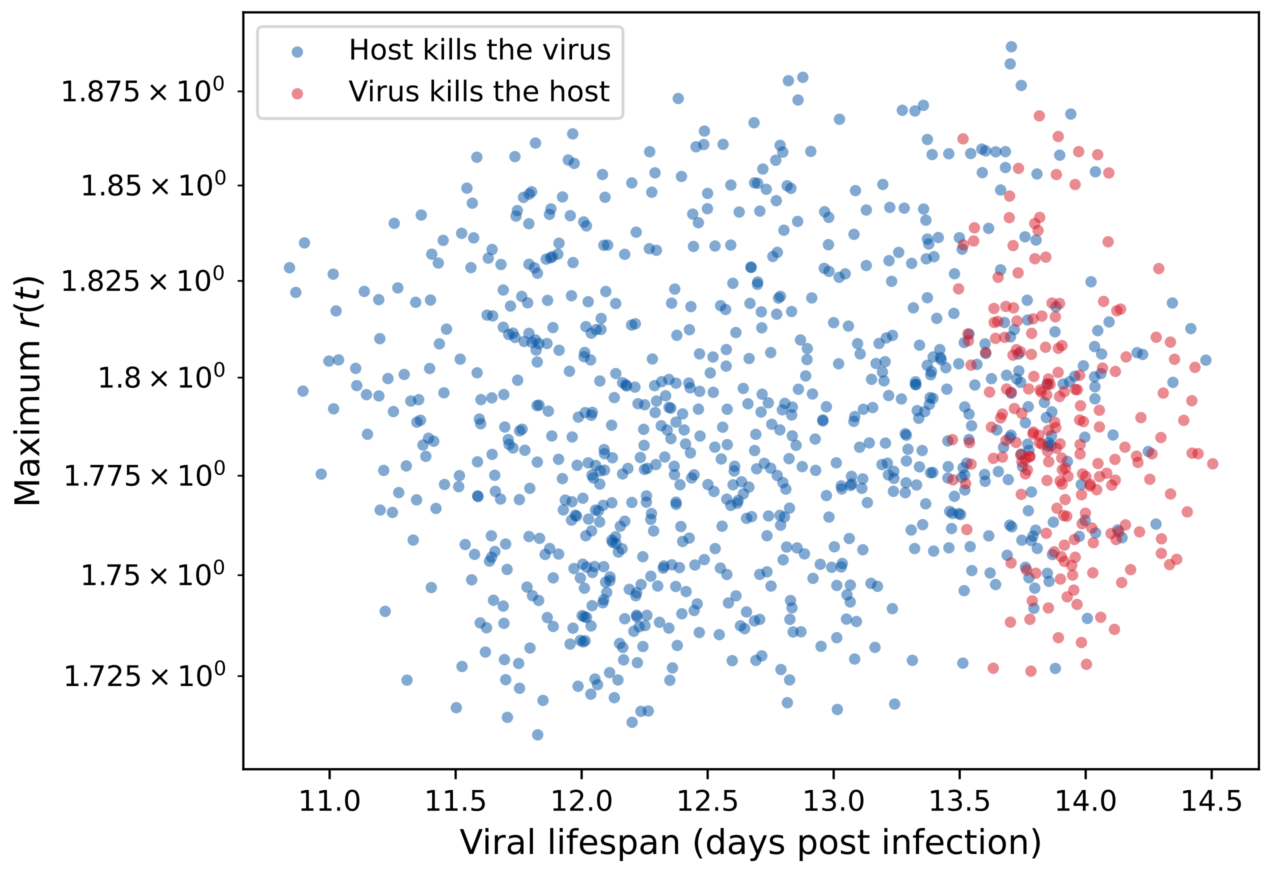


**Fig. S7.** **Maximum $r$ value vs viral lifespan assuming a lower case mortality rate (20% instead of 53%).** We found no strong correlation between maximum $r$ value and the viral lifespan. Blue circles represent H5N1 influenza patients who survive the infection (cleared the virus). Red circles represent patients who died due to the infection (where the distinction is made using the method in Section 2.1.6). Surviving individuals tended to have lower viral lifespans.

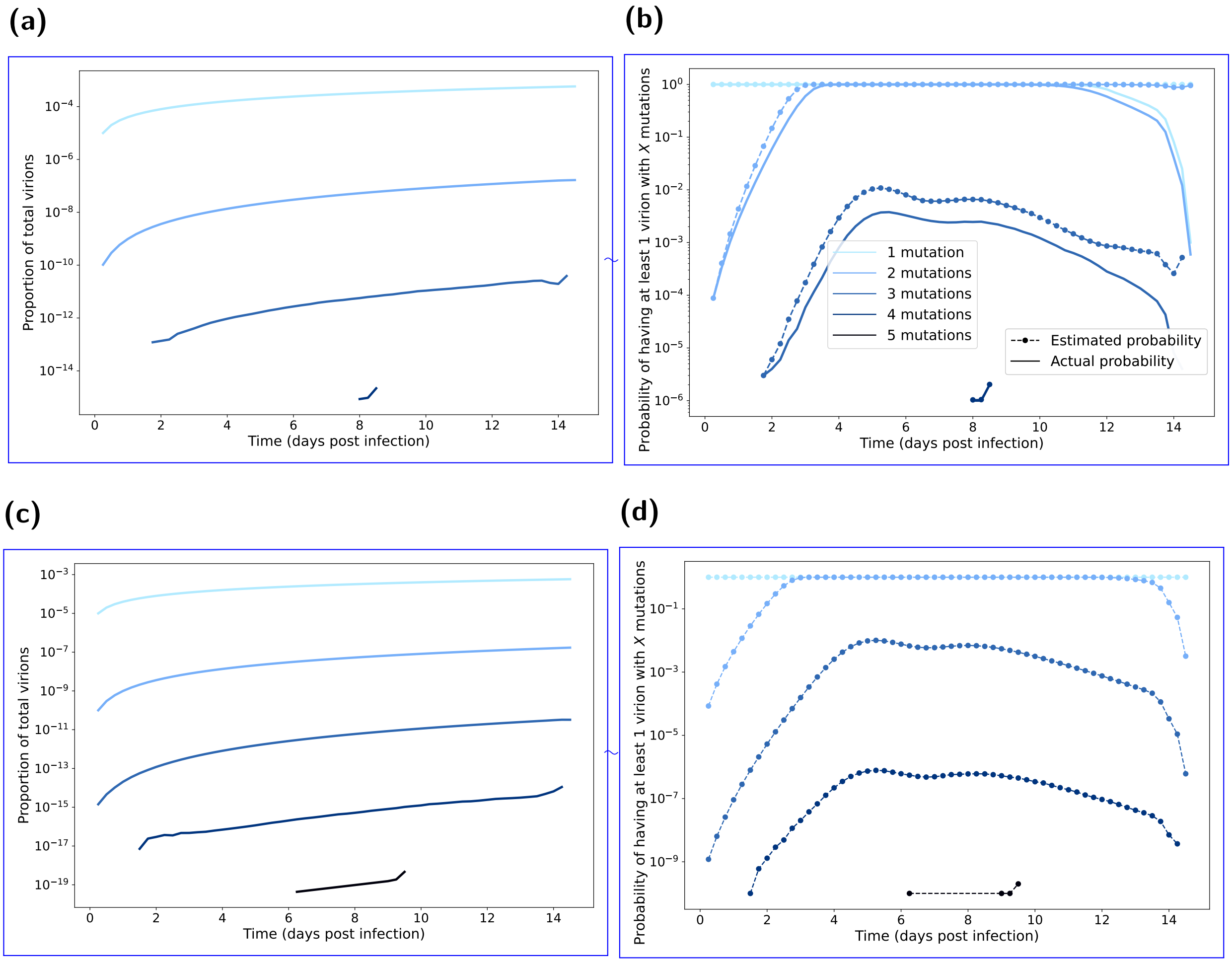


**Fig. S8.** **Mutated virion statistics with respect to time elapsed post infection, computed from BPM realisations assuming a lower case mortality rate (20% instead of 73%).** Line shading corresponds to the number of mutations (one mutation the lightest shading through to five mutations being the darkest shading). In all BPM simulations we fixed the probability of mutation at $10^{-5}$ per replication. In panels **(a&b)**, each realisation had an initial viral load of $10^6$. We ran 1000 realisations of each of the 1000 posterior parameter sets (Fig. 4). In panels **(c&d)**, each realisation had an initial viral load of $10^6 \times 10^6$. We ran one realisation of each of the 1000 posterior parameter sets (Fig. 4). **(a,c)** Proportion of total virions with the specified amount of mutations. There were a very small proportion of virions that have the required number of mutations to achieve droplet transmission (three or more mutations). **(b,d)** Probability of having a mutation strain. We present the estimated probabilities as the dashed lines with circle markers. We present the actual probabilities as solid lines. Probability estimate derivation follows that given in Section 2.2. The estimated probabilities are a clear upper bound on the true probabilities. Depending on the number of mutations in the initial infecting virions, there was a low probability of achieving the required number of mutations near the beginning on the infection lifespan (which would allow more replication of the mutant strains).
